# Use of preclinical Alzheimer’s disease trajectories for clinical trial design

**DOI:** 10.1101/2025.05.01.25326668

**Authors:** Rebecca E. Langhough, Derek L. Norton, Karly A. Cody, Lianlian Du, Erin M. Jonaitis, Rachael Wilson, Ramiro Eduardo Rea Reyes, Bruce P. Hermann, Henrik Zetterberg, Sterling C. Johnson

## Abstract

**INTRODUCTION:** This study uses longitudinal amyloid biomarker and cognitive data to generate sample size estimates for two-armed, pre-clinical amyloid clearance clinical trials.

**METHODS:** PET PiB DVR ranges defined three amyloid groups (positive, “A+”; sub threshold/low positive, “subA+”; and negative, “A-”) in cognitively unimpaired Wisconsin Registry for Alzheimer’s Prevention participants. Amyloid group trajectories estimated from mixed effects models informed per-treatment-arm sample size estimates to detect plausible treatment effects over 3-year (biomarker) or 6-year (cognition) study windows (80% power).

**RESULTS:** To detect ≥60% slowing in PiB accumulation, ≤40 may be needed per arm for both SubA+ and A+; to detect the same effect sizes in plasma p-tau217 trajectories, ∼50-1700 are needed, depending on assay and amyloid subgroup. Among cognitive outcomes, Digit Symbol Substitution and a 5-test Preclinical Alzheimer’s Cognitive Composite consistently required fewest (<2000) per arm.

**DISCUSSION:** Early intervention study planning will benefit from selection of outcomes that are most sensitive to AD biomarker-related preclinical change.

## 1 Background

As the number of people who develop Alzheimer’s disease (AD) dementia is increasing world-wide, secondary prevention clinical trials aimed at slowing or halting the disease from progressing to its symptomatic prodromal and dementia stages are of vital societal interest. Yet the preclinical stage of AD (i.e., amyloid-positive and cognitively unimpaired) may last 10-30 years(1) with only subtle evidence of cognitive decline initially (e.g., (2, 3)), such that cognitive change among A+ people in the context of a clinical prevention trial is expected to manifest as a small effect size and may require longer follow-up to detect meaningful treatment-related slowing in cognitive decline. For example, one study demonstrated that cognitive decline in baseline cognitively unimpaired (CU) participants was worse in A+ compared with A-on measures such as the Preclinical Alzheimer’s Cognitive Composite (PACC;(4)), delayed list recall, and executive function with progression from CU to MCI taking an average of 6 years in the A+ group(5).

With the advent of effective anti-amyloid therapeutics, change in amyloid burden is also a key outcome in clinical trials as this is an early detectable process that can be quantified grossly with amyloid positron emission tomography (PET; (6, 7)). While rates of biomarker and cognitive change have been studied in persons with dementia, mild cognitive impairment (MCI) and older cognitively unimpaired (CU) A+ adults, there is less known about change from earlier in the disease process when PET amyloid first becomes detectable or overtly positive. Findings from observational longitudinal studies of late midlife are needed to inform power and sample size estimates for the next generation of early intervention and prevention trials using outcomes that reflect both biological (e.g., amyloid rate of change) and cognitive responses to treatment. To maximize statistical power and ensure the trials are as efficient as possible, it is important to select outcomes that are maximally sensitive to change relative to amyloid level(8). In addition, with the recent development of blood biomarkers that are sensitive to onset of amyloid and tau abnormalities (e.g., (9–11)), blood biomarkers may be useful clinical trials outcomes in amyloid clearing trials.(12–14)

Since 2009, a subset of the Wisconsin Registry for Alzheimer’s Prevention (WRAP)(15) cohort has been participating in a longitudinal amyloid PET imaging program of research with [C-11] Pittsburgh compound B (PiB).(16–19). In 2020, we reported that rates of change in an amyloid summary index are broadly linear above the positivity threshold and increasing in a predictable manner such that both age of onset of amyloid positivity and the chronicity of the disease (defined as the time-span in years of amyloid positivity) can be feasibly estimated from a single scan.(20) We further showed in that report that preclinical cognitive decline and tau-related PET signal were more likely to be observed with longer amyloid duration. These results have been replicated by our group(21) and others(22, 23) using a variety of methods and in multiple cohorts. In addition, we have recently shown in baseline CU samples(10, 24, 25) that plasma pTau_217_ from two assays correlated well with PET amyloid and tau measures as well as with a WRAP version of the PACC outcome.(26)

The current report is a study of rates of change in PET amyloid, plasma pTau_217_ and cognition in baseline CU WRAP participants with the goal of deriving and reporting estimated slopes and sample sizes needed to detect a treatment-related reduction in amyloid rate of change and secondarily in plasma pTau_217_ and cognitive rates of change under common prevention trial scenarios for preclinical intervention. We were particularly interested in exploring rates of change in these outcomes when stratified by PET amyloid burden being in a subthreshold-to-low-positive range or an overtly positive range. Based on literature showing sex differences in cognitive outcomes and early dementia risk,(27–29) additional exploratory analyses examined if rates of change differed by sex, within each pairing of amyloid strata and outcome.

## 2 Methods

### 2.1 Cohort and Participants

WRAP, a longitudinal cohort study of persons who were non-demented at cognitive baseline (mean(sd) age=54(6) years), is enriched for AD risk via oversampling for a parental history of AD (∼72%; see Johnson et al, 2018,(15) for study design details). Briefly, WRAP participants complete comprehensive bi-annual study visits including cognitive assessments, physical exams, questionnaire completion and blood draws; a subset also complete positron emission tomography (PET) amyloid scans. At the time of these analyses, participants were included if they were CU at baseline cognitive assessment, had undergone serial cognitive assessment and had also completed at least one amyloid PET scan (n=481; see STROBE flow diagram in Supplementary Figure 1). Participants provided informed consent for all aspects of data collection, and all study procedures were conducted in accordance with the Helsinki Declaration and approved by the University of Wisconsin Institutional Review Board.

### 2.2 Neuropsychological Testing and Cognitive Outcomes

The WRAP neuropsychological assessment battery has been described in detail elsewhere.(15) The test battery includes tests that represent multiple cognitive domains and comprehensively, are sensitive to cognitive change throughout the cognitive performance continuum (i.e., from CU to dementia). For these analyses, we selected individual tests and composites created using them that have been shown to be sensitive to preclinical cognitive decline. Individual tests included the Rey AVLT sum of learning trials (AVLT; (30)); Digit Symbol Substitution Test (31), Trail making task-Trial A and Trial B (TMT-A, TMT-B;(32)), Logical Memory delayed recall (LMT-II; (33)); phonemic fluency task using letters C, F and L (CFL; (34)); and semantic fluency animal naming task (Animal Naming; (35)).

Composites included the following: two WRAP versions of the Preclinical Alzheimer’s Cognitive Composite(26) and a processing speed (PS) composite. The three-test PACC (PACC3) was calculated using the AVLT sum of learning trials, Digit Symbol substitution, and Logical Memory delayed recall. The PACC5 included those tests plus CFL and TMT-B time. The PS composite was calculated as the average of Digit Symbol Substitution Test and Trail making task-Trial A (TMT-A) z-scores based on findings in the Harvard Aging Brain Study (HABS) showing that these two tests were declining in participants in early stages of amyloid accumulation (i.e., <20 Centiloid; (36)). Prior to calculating composites, individual tests were converted to z-scores using the mean(sd) of baseline CU participants; tests where higher raw scores indicate worse performance were reflected so that higher indicated better performance across all z-scores. Composites were then calculated as the average of contributing z-scores.

Given the significant role that study partner reports play in staging clinical dementia severity at more advanced stages of decline, we also included reports of functioning provided by study partners in these analyses. Specifically, in 2012, WRAP began administering the Clinical Dementia Rating Scale (CDR; (37)), a validated interview-based system to stage dementia severity. The CDR yields a global score (0=no impairment; .5, 1, 2, and 3 representing increasing levels of impairment) and a sum of boxes (SB) score based on ratings for six domains of functioning (CDR-SB possible range=0-18 with scores between 0.5 and 4 indicating questionable cognitive impairment to potentially very mild dementia;(38)). Later, WRAP began using the Quick Dementia Rating System (QDRS; (39)) as an initial screen. The QDRS takes only 3-5 minutes to complete, yields multiple scores, including a QDRS total score that includes all ten items (possible range 0-30), as well asscores parallel to the CDR global and CDR-SB.(40, 41) For sample size projections, we used the following three study partner-based variables: harmonized QDRS/CDR-SB (based on the six items similar on both study partner measures and using CDR item scores, if available, and QDRS item scores if there was no CDR); QDRS-SB (using the six items that contributed to the harmonized QDRS/CDR-SB); and QDRS Total.

### 2.3 Cognitive Status Determination

Participant cognitive status in WRAP is assigned after each study visit using a two-tiered consensus review process described in detail elsewhere.(42) Briefly, if participant performance is flagged as potentially abnormal based on data from the neuropsychological assessment visit or informant reports, the participant’s cognitive status is determined by complete review of the participant’s data by a multidisciplinary consensus team. The team assesses whether participants meet clinical criteria based on internal norms, raw scores, or informant reports (Mild Cognitive Impairment (MCI) criteria as in (43) and dementia criteria as in (44). For those who do not meet criteria for MCI or dementia, a status of “Cognitively Unimpaired-Declining” (CU-D) is assigned when cognitive performance is lower than expected, but not low enough for an MCI or dementia diagnosis; those with little or no evidence of mild deficits are labeled as “Cognitively Unimpaired-Stable” (CU-S). For these analyses, CU-S and CU-D were combined into a single CU group.

### 2.4 PET amyloid assessment

PET amyloid burden was quantified from a global cortical composite of eight bilateral ROI’s using the tracer 11C-Pittsburgh Compound B to obtain a global PiB DVR from each scan as described in (45). A group-based trajectory modeling (GBTM) approach was applied to global PiB DVR to estimate amyloid onset age (EAOA) and PiB DVR values at the cognitive and plasma assessments (see (20) and (21) for modeling details). EAOA was the estimated age at which a person had a global PiB DVR of >=1.20; the cut-off was used in the original amyloid onset modeling paper (20) and was based on a prior ROC comparison with visual read.(19) This DVR A+ threshold corresponds roughly to 23 on the Centiloid (CL) scale, a standardized scale to estimate brain amyloid across multiple tracers.(46)

### 2.5 Plasma p-tau217 assays

Plasma sampling methods have been described in detail elsewhere (24). In these analyses, we included plasma p-tau217 values from two assays available on subsets of WRAP EDTA plasma at the time of these analyses. Specifically, longitudinal plasma p-tau217 was assayed in a PET subset using the Meso Scale Discovery platform for 158 participants in this sample (504 total visits; pTau217_Meso;_ (24)) and the Quanterix ALZpath Single molecule array (SIMOA®) assay for 336 WRAP participants in this sample (1159 total visits; pTau217_ALZpath_; (47)).

### 2.6 Statistical Analysis

#### 2.6.1 General consideration

All analyses were performed in R version 4.4.0(48) with all mixed effects models fit using the *lme4*(49) package, descriptive statistics of the amyloid groups tabulated with the *tableone*(50) package, and graphics produced with the *ggplot2*(51) package. Partitioning the samples into amyloid-level estimation groups.

For primary analyses to estimate slopes needed for subsequent sample size calculations, visit-level data for each outcome were partitioned into three amyloid groups based on observed PiB global DVR (for PiB-based sample size estimates) or modeled/imputed PiB global DVR at the time of a cognitive assessment study visit for outcomes obtained asynchronously from PiB scans (for details on this estimation, see(21) supplementary materials). The amyloid groups included amyloid negative (“A-“: global PiB DVR<1.13) sub-threshold-to-low-amyloid-positive (“subA+”; DVR in[1.13, 1.2); and amyloid-positive (“A+”; Global PiB DVR≥1.2). The lower and upper cut-offs of 1.13 and 1.20 correspond approximately to Centiloid values of 12.7 and 23.0 using a published linear transformation of local global PiB DVR values to the Centiloid scale (21). The lower cut-off corresponds to literature estimating CL 12-13 as a disease onset inflection point from non-accumulating to amyloid accumulating (e.g,(52, 53)) while the upper cut-off is consistent with CL values representing overtly amyloid positive levels (e.g., (54, 55)). The middle group includes the CL range of 15-18.5 recently identify by Farrell and colleagues as “the lowest point in a baseline distribution that robustly predicts future Aβ accumulation and cognitive decline”(56) and thus is labeled sub-to-low A+ (subA+).

The above process was used to maximally leverage the data available for estimation of placebo/control group trajectory and general error terms while attempting to reasonably mimic a clinical trial setup with an observational dataset. While many subjects have well over 6 years of data, only up to 3 or 6 years of data was used (depending upon whether the outcome was a biomarker or a cognitive outcome) for a given estimation group to represent the timeframe of the theoretical clinical trial. In addition, this partitioning approach results in many subjects contributing data to not just one, but two or all three estimation groups (A-, subA+, A+), with some data points being used in multiple groups, and other data points only being used in one group. However, in any one estimation group, only up to 3 or 6 years of visits were used, and thus most subjects do not contribute all of their data to any one given estimation group, and some data on subjects are not used at all if consecutive visits span too much time (i.e., 3 years for biomarkers and 6 years for cognitive outcomes). Figures 1A and 1B illustrate this partitioning for three participants each for PiB and cognition, respectively; in each set of three, two participants’ longitudinal trajectories within the study supported their inclusion in more than one group. In exploratory analyses, we further partitioned the A+ group into A+_<40CL_ for raw or estimated global PiB DVR [1.2, 1.32) and A+_>=40CL_ for raw or estimated global PiB DVR >= 1.32 (∼>=40 CL) as these ranges were similar to recently conducted clinical trials of people with MCI or mild dementia (e.g., (7)).

**Figure 1:**
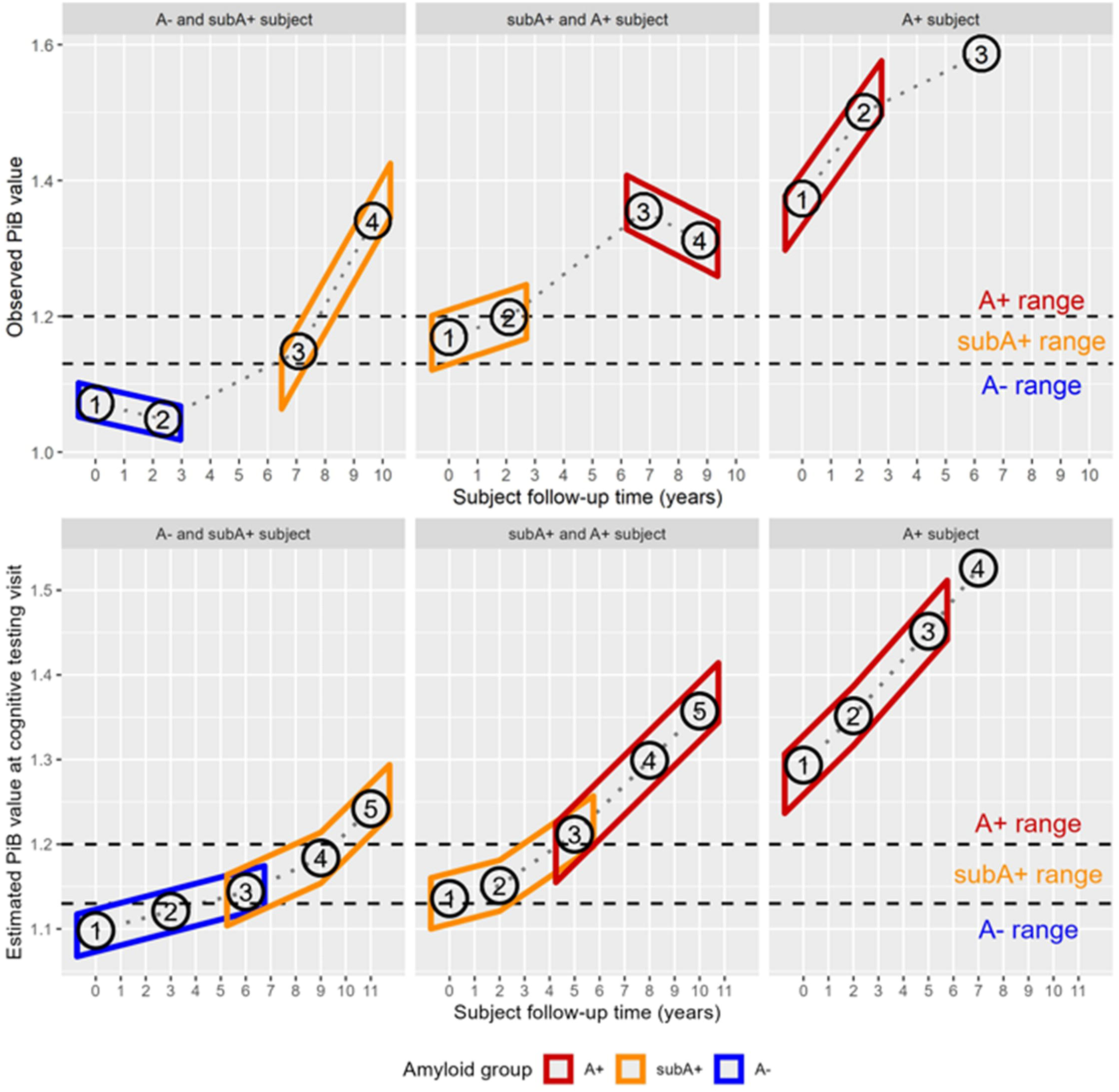
Illustration of partitioning of participant visits for slope estimates. **In panel A** (top), partitioning of PiB DVR data is shown for three participants. The participant at the top left transitioned from A- to A+ between PiB visits 2 and 3, which were 2.5 and 7 years post baseline PiB scan, respectively. The first two visits met criteria for slope estimates in the A-group while the second two visits contributed to the slope estimates for the subA+ group. In the top middle panel, the participant had two visits each that contributed to slope estimates in the subA+ and A+ groups. In the panel at the top right, the participant’s first two visits contributed to slope estimates for the A+ range, while the third visit was not included because it was beyond the trial design window of 3 years for PiB. **Panel B** (bottom) shows partitioning of cognitive visit data based on the imputed/modeled PiB DVR at each cognitive visit for the same participants to estimate slopes for a six-year cognitive follow-up window. Abbreviations: A-= amyloid negative group (CL<∼13); subA+ = sub-threshold to low positive group (CL ∼13-23); and A+ = overtly A+ group (CL >∼23); DVR=Distribution Volume Ratio; PiB= Pittsburgh Compound B.

Visit-level data were included in slope estimates for each amyloid group using the following process. For each participant, each visit was examined to determine whether the corresponding observed or imputed DVR qualified as an “initial visit” in one of the amyloid groups, where “initial visit” was the first available (i.e., youngest age) visit that was in the amyloid group’s range. In addition, for a visit to qualify as an initial visit in a particular amyloid group, the participant had to be between 50-85 years and cognitively unimpaired (CU) at that visit. Finally, to be included in slope estimations for that amyloid group, the participant had to have had at least two visits for the outcome of interest within 3 years for biomarkers and 6 years for cognition. For each outcome, all available visits within 3 years (biomarkers) or 6 years (cognitive outcomes) from this “initial visit” were then used in slope estimates for that amyloid group. We selected these clinical trial design time-frames (3 or 6 years) based on studies showing a need for longer studies when biomarker burdens are lower and cognitive change is slower.(5, 36)

#### 2.6.2 Estimating outcome slopes within each group

After partitioning the data into amyloid groups, linear mixed effects models provided amyloid group-specific slope and error estimates for Global PiB DVR, plasma pTau217 (two assays) and the previously mentioned cognitive composites and individual tests considered sensitive to AD-related change. Models used fixed effects of participant years since the first visit used in the amyloid grouping, and random effect of participant-specific intercepts; participant-specific random slopes were not used due to instability issues in some of the amyloid group / outcome pairings with smaller sample sizes, and the use of uniform methods across all the outcomes analyzed. No other terms were included in primary slope estimation models.

#### 2.6.3 Sample size estimations

The fixed effects slope estimates and residual errors from 2.6.3 models were then used to estimate sample sizes needed to achieve 80% power in a typical double-blinded, two-arm clinical trials of participants who were considered to be subA+ or A+ and CU at trial enrollment. We estimated sample sizes relative to multiple theoretical treatment effect sizes in the treated arm using higher expected treatment effect sizes for biomarkers (i.e., 60-90% reduction in amyloid, 20-35% reduction in cognitive decline). As noted earlier, PiB and plasma amyloid outcome scenarios used a 3 year follow-up time and estimation process (mimicking a 3 year clinical trial time) for these sample sizes, and the cognitive tests and composites used a 6-year time period. For primary analyses, treatment effects were calculated as percent change relative to slope estimates within the SubA+ and A+ groups. That is, the observed change within an amyloid group was used to estimate mean change over the trial duration in the trial’s control group and the estimated treatment group change was reduced by the effect sizes of interest (i.e., and is referred to as “attenuation towards 0” or typical decline for this group in the absence of intervention). Error for the biomarker or cognitive outcomes at trial entry and end was taken as the residual error from the fitted model for the associated outcome (resulting in an error scaled by the square-root of 2 for a paired difference). All models / sample sizes were calculated on the outcomes’ raw scales, except for TMT-A and TMT-B which were log transformed before being fitted / sample sizes calculated, due to heteroskedasticity issues of the fitted models on the raw scale. Sample sizes were then calculated from a two-tailed, two-sample t-test set up using these estimated means and errors for treatment and control groups and the aforementioned effect size ranges (80% power).

#### 2.6.4 Exploratory/sensitivity analyses

2.6.4.1 The “attenuation to 0” estimates assume that slopes in the A-group are flat. To illustrate how not accounting for “normal aging” declining slopes might impact sample size calculations, we also calculated sample sizes for treatment effects *relative to the A-slopes* for the SubA+ group (representing attenuation to normal preclinical age-related change in the treatment group). Especially for the treatment effect on the SubA+ group, targeting a slope of 0 could represent an overly optimistic treatment effect, especially if the A-groups slope is closer to SubA+ group slope than to 0. Equations 1 and 2 below show how the subA+ treatment group slope was estimated for attenuation to 0 (Equation 1) and to the A-slope (Equation 2), respectively:

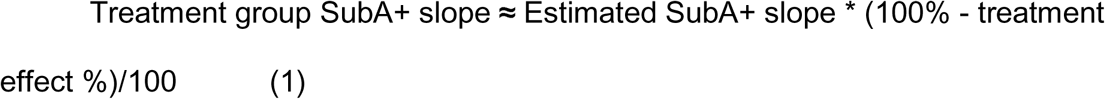

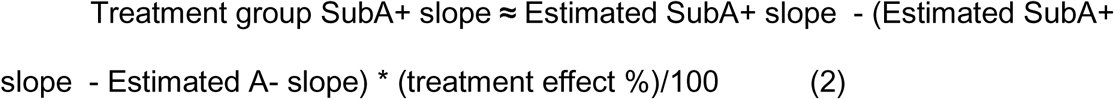

2.6.4.2 In additional exploratory analyses based on clinical trials targeting specific ranges such as 20-40 CL (e.g., (7)), we repeated the partitioning of data, slope estimation, and sample size estimation steps of 2.6.1-2.6.3 for the A+ subgroups split at 40 CL (i.e., A+_<40CL_ and A+_>=40CL_).

2.6.4.3 With respect to planning a clinical trial, if there is a substantial difference between men and women in cognitive or biomarker trajectories, this could lead to trial designs needing to stratify by sex before randomization and could result in different enrollment numbers for each sex to achieve similar powers for the same treatment effect. Therefore, as a sensitivity analysis related to slope estimates, we examined if there is evidence for a sex difference in the trajectory data for the various outcomes and amyloid groups described above by including a sex indicator interacting with time in the characterizing regression models and testing for significance.

## 3 Results

### 3.1 Biomarker outcomes

Sample characteristics for biomarker subsets (by outcome type and amyloid estimation group) are shown in Table 1 for amyloid PET and supplementary Table 1 for the two plasma p-tau217 biomarkers. *APOE□*4 count patterns follow that expected across the estimation groups, with those in the A-group most likely to have 0□4 alleles (∼64-68% of the A-group, 41-50% of subA+’s, and 27-31% of A+’s having 0□4 alleles), and relatively higher proportions in the A+ group carrying 2□4 alleles (1-2% of the A-group, 0-10% in subA+ group, and 14-20% in the A+ group). The A-group’s average age is about three years younger than the average age in the subA+ and A+ group. Observed longitudinal PiB, pTau217_Meso_, and pTau217_ALZpath_ spaghetti plots are shown vs age in Supplementary Figure 2. In Supplementary Figure 3, the two pTau217 outcomes are each shown vs estimated PiB DVR at plasma draw. For both figures, color coding denotes amyloid estimation groups.

**Table 1.** Sample Characteristics by Biomarker outcome and amyloid groupse.

|  |  |  |  |  |
| --- | --- | --- | --- | --- |
|  | Amyloid Groups |  |  | Total in calculations |
|  | A- | subA+ | A+ |  |
| **Summary information for PiB DVR estimation sample** |  |  |  |  |
| Number of participants per amyloid group with $\geq 2$ PiB DVRs | 151 | 16 | 40 | 197 |
| Number of observations per group | 302 | 32 | 80 | 414 |
| Baseline amyloid PET Scan Age, median [min, max] | 61.7 [50.0, 79.2] | 64.6 [60.6, 71.9] | 66.4 [52.4, 79.0] | 62.7 [50.0, 79.2] |
| Self-reported Sex, n(%) |  |  |  |  |
| Female | 102 (67.5) | 12 ( 75.0) | 23 (57.5) | 131 (66.5) |
| Male | 49 (32.5) | 4 ( 25.0) | 17 (42.5) | 66 (33.5) |
| Race/ethnicity, n(%) non-Hispanic White/Caucasian |  |  |  |  |
| non-Hispanic White/Caucasian | 140 (92.7) | 16 (100.0) | 38 (95.0) | 184 (93.4) |
| Hispanic or non-White/Caucasian | 11 ( 7.3) | 0 ( 0.0) | 2 ( 5.0) | 13 ( 6.6) |
| missing | 0 ( 0.0) | 0 ( 0.0) | 0 ( 0.0) | 0 ( 0.0) |
| APOE carrier status, n(%) |  |  |  |  |
| 0 ε4 alleles | 97 (64.2) | 8 ( 50.0) | 11 (27.5) | 112 (56.9) |
| 1 ε4 allele | 48 (31.8) | 8 ( 50.0) | 23 (57.5) | 73 (37.1) |
| 2 ε4 alleles | 4 ( 2.6) | 0 ( 0.0) | 6 (15.0) | 10 ( 5.1) |
| missing | 2 ( 1.3) | 0 ( 0.0) | 0 ( 0.0) | 2 ( 1.0) |
| Education, n(%) with college degree or higher |  |  |  |  |
| college degree or higher | 104 (68.9) | 12 ( 75.0) | 31 (77.5) | 140 (71.1) |
| high school educated or less | 44 (29.1) | 4 ( 25.0) | 9 (22.5) | 54 (27.4) |
| missing | 3 ( 2.0) | 0 ( 0.0) | 0 ( 0.0) | 3 ( 1.5) |
| WRAT reading standard score, mean(std) | 106.53 (9.92) | 109.12 (7.54) | 106.92 (7.37) | 106.76 (9.39) |
| Cognitive Status at last cognitive assessment, n(%) |  |  |  |  |
| CU-S | 119 (79.3) | 15 ( 93.8) | 31 (77.5) | 156 (79.6) |
| CU-D | 31 (20.7) | 1 ( 6.2) | 6 (15.0) | 37 (18.9) |
| Impaired (not MCI) | 0 ( 0.0) | 0 ( 0.0) | 0 ( 0.0) | 0 ( 0.0) |
| MCI | 0 ( 0.0) | 0 ( 0.0) | 3 ( 7.5) | 3 ( 1.5) |
| Dementia | 0 ( 0.0) | 0 ( 0.0) | 0 ( 0.0) | 0 ( 0.0) |
| Note: Partitioning of participant visit-level data is described in section 2.6.2. |  |  |  |  |
| Abbreviations: A- = amyloid negative group (CL<~13); subA+ = sub-threshold to low positive group (CL ~13-23); and A+ = overtly A+ group (CL >~23); APOE=apolipoprotein E; CU=Cognitively Unimpaired (subtypes: S=Stable; D=Declining); DVR=Distribution Volume Ratio; MCI=mild cognitive impairment; PET=Positron Emission Tomography; PiB=[C-11]Pittsburgh Compound B; WRAT=Wide Range Achievement Test |  |  |  |  |

Figure 2 depicts the 95% CI’s for average estimated biomarker slopes by A-, subA+, and A+ amyloid estimation groups. Slopes and CI’s for raw values are depicted in the top row while standardized estimates are depicted in the bottom row to facilitate visual comparison across biomarkers. Slopes did *not* differ from zero (p>.05; green color) for PiB DVR and plasma pTau217_Meso_ in the A-group and for pTau217_ALZpath_ in the subA+ group. Slopes were positive (p<.05) for the remaining subA+ and A+ estimates. Supplementary Figure 4 shows the parallel version with the A+ group partitioned into A+_<40CL_ and A+_>=40CL.._ For both plasma markers, slope estimates are higher in the A+_>=40CL_ group compared to the A+_<40CL_ group. The slopes(standard errors) and contributing sample sizes for the biomarker outcomes are shown in the biomarker section of Supplementary Table 2.)

**Figure 2:**
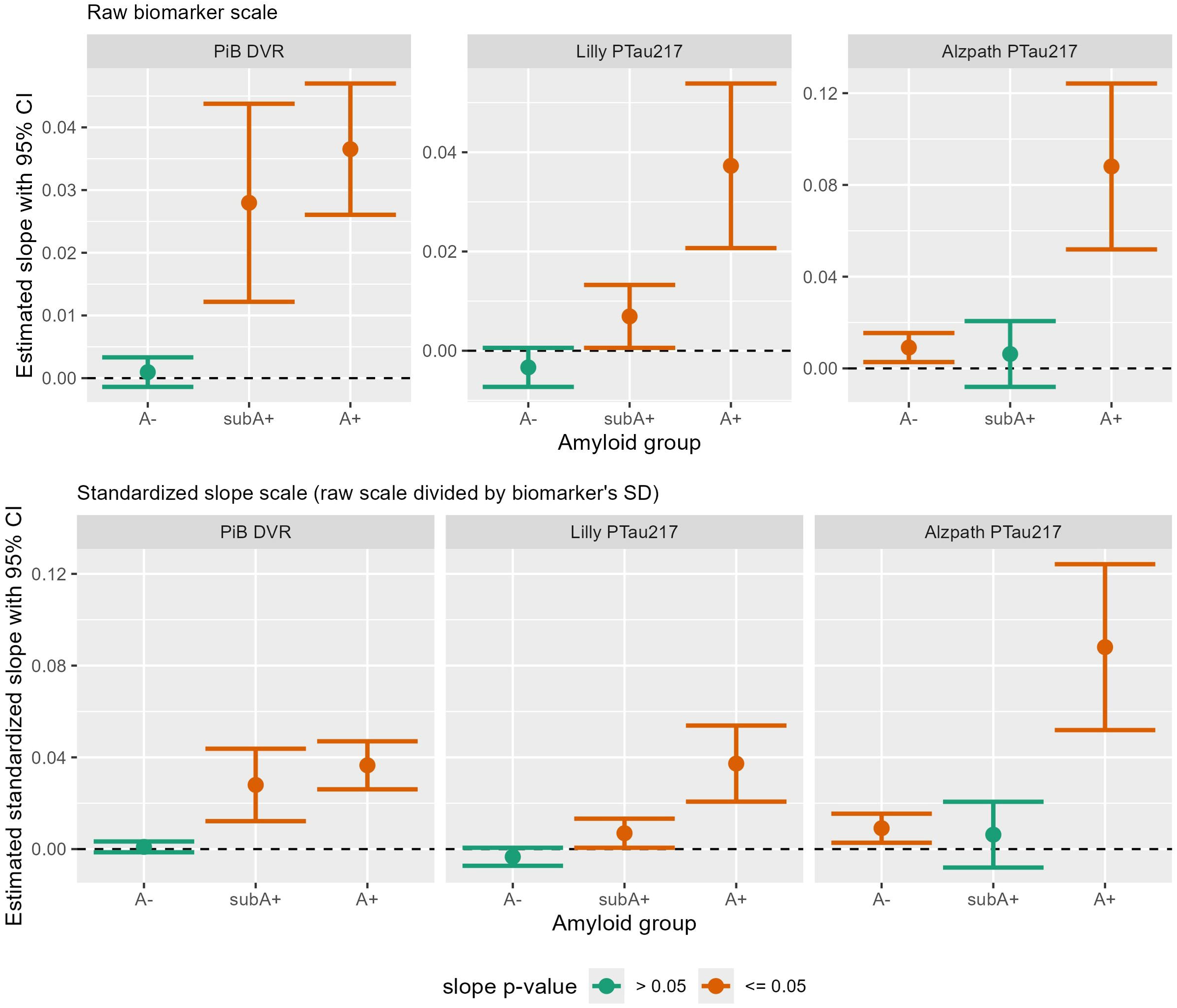
Estimated 3-year regression slopes with 95% CI’s of biomarkers by amyloid group. The top row depicts estimated slopes by amyloid group for PiB DVR (left), Lilly MSD plasma pTau217 pg/mL (middle), and Quanterix ALZpath plasma pTau217 pg/mL (right). The bottom row shows the same data using standardized slope estimates to facilitate visual comparisons of effect sizes across biomarkers. Slopes that differ significantly from zero are in orange while those not differing from zero are in green. Abbreviations: A-= amyloid negative group (CL<∼13); subA+ = sub-threshold to low positive group (CL ∼13-23); and A+ = overtly A+ group (CL >∼23); CL=Centiloid; DVR=Distribution Volume Ratio; Lilly MSD= Lilly Meso Scale Discovery; PiB= Pittsburgh Compound B; pTau217= phosphorylated tau protein at amino acid 217.

Figure 3 depicts sample size estimates to have 80% power to detect treatment effect sizes of 60-90% reduction in biomarker accumulation/worsening using the slope(error) estimates from Figure 2 and the two-sample paired t-test approach outlined in 2.6.4. In the subA+ group, sample size estimates for the PiB DVR outcome ranged from 37 participants per treatment arm for 60% attenuation to zero, to 17 participants per arm for 90% attenuation to 0 (Figure 3, red line in top left panel). Corresponding estimates for 60 to 90% attenuation to the A-slope were slightly higher and ranged from 39 down to 18 per treatment arm, respectively (Figure 3, red dotted line in top right panel). Similarly, in the A+ group, 21 or 10 per treatment arm would be needed to have power to detect 60% or 90% reduction in PiB DVR accumulation, respectively (Figure 3 red line in bottom left panel). Estimated sample sizes for the same treatment effects ranged from 12 down to 6 for A+_<40CL_ (Figure 3, red line, triangles in bottom right panel), and from 25 down to 12 for A+_>=40CL_ (Figure 3: red line, squares in bottom right panel). Sample size requirement estimates were substantially higher for the two plasma pTau217 markers for both the subA+ and A+ groups, as noted below (Sample size estimates for 60%-90% attenuation in worsening are also listed in Supplementary Table 3 and depicted with greater precision in Supplementary Figure 5.)

**Figure 3:**
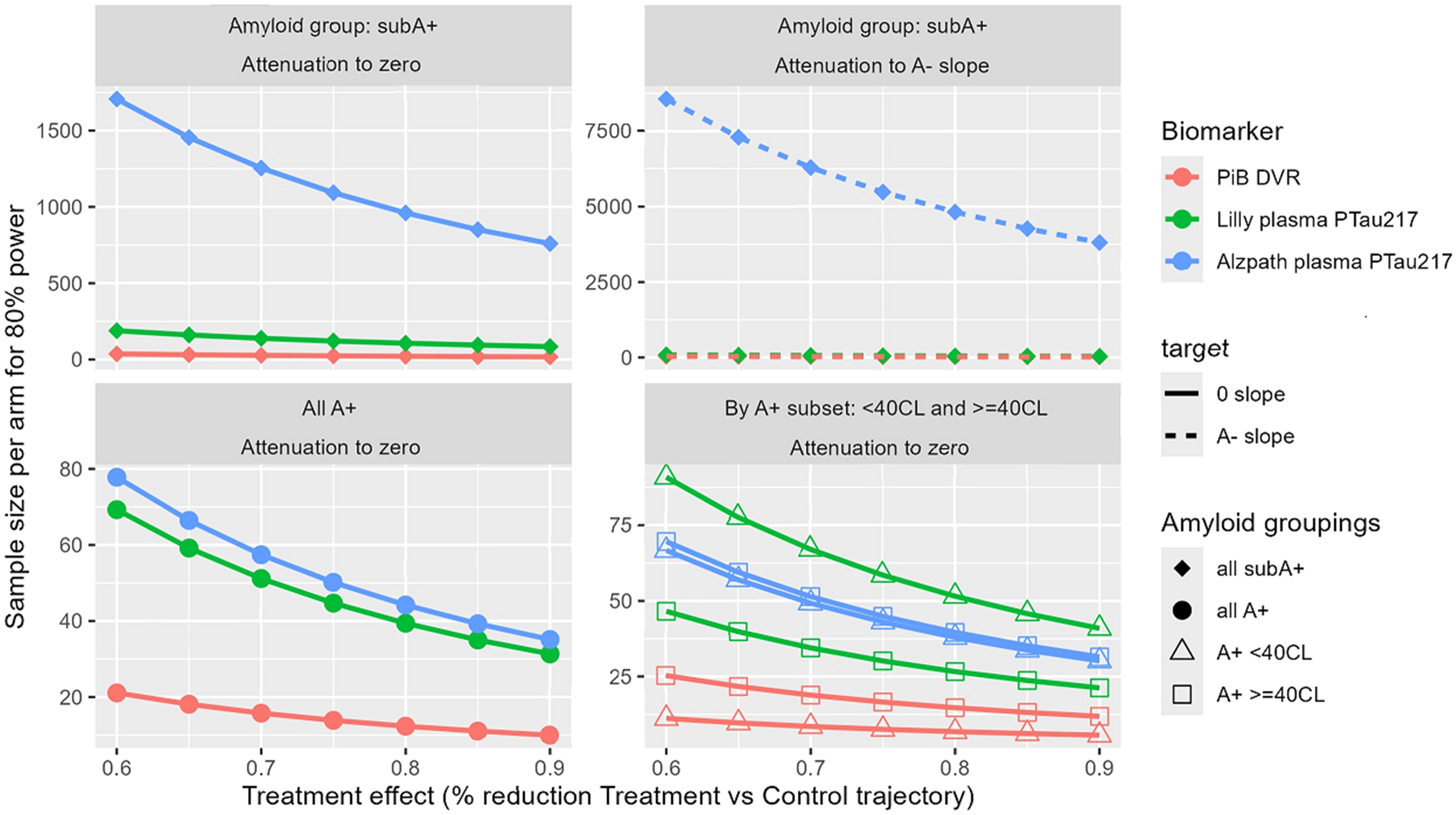
Sample size projections for biomarker outcomes. Sample size projections for biomarker outcomes. Red, green, and blue indicate numbers of participants needed per treatment arm to have 80% power to detect effect sizes of 60-90% reduction in treatment vs control trajectories for PiB DVR, Lilly MSD pTau217, and ALZpath pTau217, respectively. N’s are shown for studies targeting amyloid clearance in the subA+ group (top row: left panel depicts slope attenuation to 0 while right panel depicts slope attenuation to the A-group) and in the A+ group (bottom row: left panel depicts slope attenuation to 0 and right panel shows sample size needed within low A+ and higher A+ subgroups.) Sample size estimates are also listed in Supplementary Table 3. Abbreviations: A-= amyloid negative group (CL<∼13); subA+ = sub-threshold to low positive group (CL ∼13-23); and A+ = overtly A+ group (CL >∼23); CL= Centiloid; DVR=Distribution Volume Ratio; Lilly MSD= Lilly Meso Scale Discovery; PiB= Pittsburgh Compound B; pTau217= phosphorylated tau protein at amino acid 217.

#### pTau217_Meso_

In the subA+ group, 60 and 90% attenuation to 0 in pTau217_Meso_ change would require 189 to 85 per treatment arm (Figure 3, green line in upper left panel) while the same treatment effects based on attenuation to the A-slope would require fewer per treatment arm (87-39; Figure 3, green dotted line in upper right panel). In the A+ group, 70 to 32 per treatment arm would be needed to have power to detect 60% or 90% reduction in plasma p-tau217 change, respectively (Figure 3 green line lower left panel). Estimates for the same treatment effects were 91 and 41 for A+_<40CL_ (Figure 3 green line, triangles, lower right panel) and 47 to 22 for A+_>=40CL_ (Figure 3 green line, squares, lower right panel). See also Supplementary Table 3 and Supplementary Figure 5 for details.

#### pTau217_ALZpath_

In the subA+ group, the low p-tau217 slope resulted in very high sample size estimates relative to the other two biomarkers. Specifically, to have adequate power to detect 60 and 90% attenuation to 0 in pTau217_ALZpath_ change would require 1708 to 760 per treatment arm (Figure 3: blue line upper left panel). Since the A-slope is slightly higher than the subA+ slope for this assay, it does not make sense to report sample sizes needed for the same treatment effects based on attenuation to the A-slope. In the A+ group, 78 to 36 per treatment arm would be needed to have power to detect 60% or 90% reduction in plasma p-tau217 change, respectively (Figure 3: blue line lower left panel). Estimates for the same treatment effects were 67 and 31 for A+_<40CL_ (Figure 3: dotted blue line, triangles) and 70 to 32 for A+_>=40CL_ (Figure 3: dotted blue line, squares). See also Supplementary Table 3 for sample sizes to detect 60, 70, 80, and 90% reduction in slopes in the treatment group.

### 3.2 Cognitive outcomes

Sample characteristics for cognitive outcomes are shown in Table 2 by amyloid-level estimation group. Patterns in demographics are similar to those seen in Table 1. Average baseline scores within each amyloid estimation group are shown in Supplementary Table 4. Figure 4 depicts the 95% CI’s for average estimated cognitive slopes by A-, subA+, and A+ estimation groups for the three composites and the eight individual cognitive tests. Slopes in the CU, A-group showed significant decline for PACC3 and Digit Symbol Substitution; the rest of the slope estimates in the A-showed either positive slopes (CFL) or did not differ from 0 (the rest of the cognitive outcomes). In the subA+ group, slopes indicated significant decline on all three composites and on the Digit Symbol Substitution test; the latter had the largest standardized effect size (see Supplementary Figure 6 for depiction of standardized slopes). In the A+ group, both PACC composites, both AVLT outcomes, Delayed Story Recall and Digit Symbol Substitution tests showed significant preclinical decline while the remaining tests did not (PS composite, both Trails, CFL and Animal Naming). Standardized slopes indicate greatest A+ group declines in Digit Symbol Substitution, followed by the PACC composites. In the A+ subgroups, results generally showed slightly greater decline in the A+_>=40CL_ subgroup (Supplementary Figure 7). The slopes(standard errors) and contributing sample sizes for the cognitive outcomes are shown in the middle section of Supplementary Table 2.)

**Figure 4:**
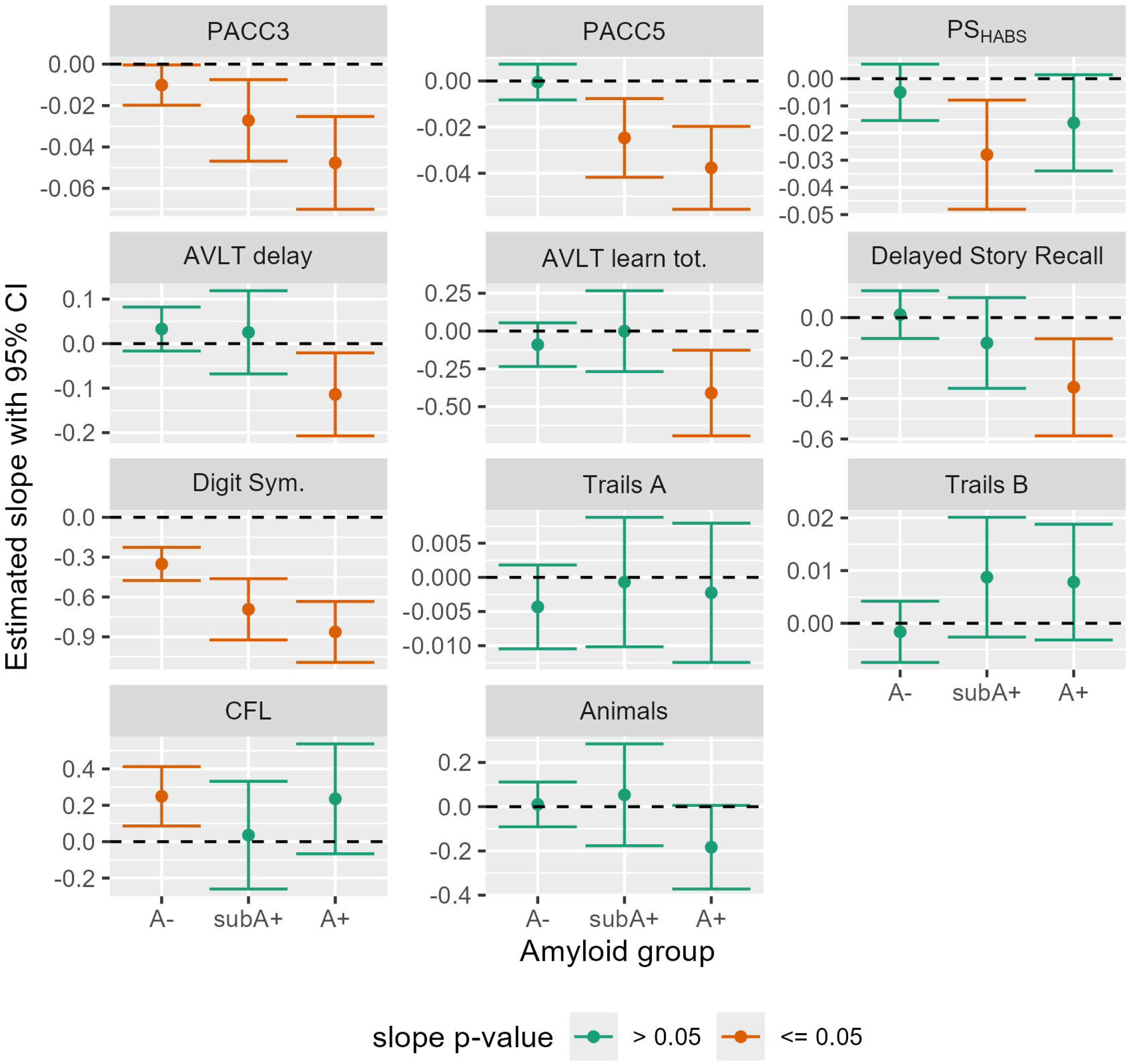
Estimated 6-year regression slopes with 95% CI’s of cognitive composite and individual test by amyloid group. Estimated slopes (test change direction) with 95% confidence intervals for cognitive composites (top row) and individual tests (bottom 3 rows), by amyloid group. Green indicates slope did not differ from 0 (p>0.05) while orange indicates slopes that differed from 0. For all but Trails A and B, negative numbers indicate decline. Abbreviations: A-= amyloid negative group (CL<∼13); subA+ = sub-threshold to low positive group (CL ∼13-23); and A+ = overtly A+ group (CL >∼23); CI=Confidence interval; CL=Centiloid; Animals: Semantic fluency with animal naming; AVLT: Rey Auditory Verbal Learning Test; Digit Sym: CFL: Phonemic fluency with letters C, F, and L; CI= Confidence Interval; Digit Sym.=Digit Symbol Substitution Test; PACC: Preclinical Alzheimer’s Cognitive Composite (3 and 5 test versions); PS: Processing Speed composite (inspired by Harvard Aging Brain Study (HABS) composite).

**Table 2.** Demographics’ summary for cognitive outcomes.

| Table 2 - Demographics' summary for cognitive outcomes |  |  |  |  |
| --- | --- | --- | --- | --- |
| Summary information for cognition estimation sample | A- | subA+ | A+ | Total in calcs |
| Number of participants per amyloid group with ge 2 with cognitive timepoints* | 382 | 109 | 92 | 439 |
| Number of observations per group | 1973 | 336 | 381 | 2437 |
| Baseline Cognitive Assessment Age, median [min, max] | 54.0 [50.0, 73.0] | 62.0 [50.0, 79.0] | 62.0 [50.0, 74.0] | 55.0 [50.0, 73.0] |
| Self-reported Sex, n(%) |  |  |  |  |
| Female | 262 (68.6) | 79 (72.5) | 68 (73.9) | 304 (69.2) |
| Male | 120 (31.4) | 30 (27.5) | 24 (26.1) | 135 (30.8) |
| missing | 0 ( 0.0) | 0 ( 0.0) | 0 ( 0.0) | 0 ( 0.0) |
| Race/ethnicity, n(%) non-Hispanic White/Caucasian |  |  |  |  |
| non-Hispanic White/Caucasian | 360 (94.2) | 105 (96.3) | 89 (96.7) | 414 (94.3) |
| Hispanic or non-White/Caucasian | 22 ( 5.8) | 4 ( 3.7) | 3 ( 3.3) | 25 ( 5.7) |
| missing | 0 ( 0.0) | 0 ( 0.0) | 0 ( 0.0) | 0 ( 0.0) |
| APOE carrier status, n(%) |  |  |  |  |
| 0 ε4 alleles | 246 (64.4) | 46 (42.2) | 30 (32.6) | 262 (59.7) |
| 1 ε4 allele | 118 (30.9) | 51 (46.8) | 45 (48.9) | 146 (33.3) |
| 2 ε4 alleles | 12 ( 3.1) | 11 (10.1) | 17 (18.5) | 24 ( 5.5) |
| missing | 6 ( 1.6) | 1 ( 0.9) | 0 ( 0.0) | 7 ( 1.6) |
| Education, n(%) with college degree or higher |  |  |  |  |
| college degree or higher | 255 (66.8) | 73 (67.0) | 64 (69.6) | 297 (67.7) |
| high school educated or less | 122 (31.9) | 35 (32.1) | 27 (29.3) | 137 (31.2) |
| missing | 5 ( 1.3) | 1 ( 0.9) | 1 ( 1.1) | 5 ( 1.1) |
| WRAT reading standard score, mean(std) | 106.46 (9.46) | 107.06 (8.98) | 106.46 (9.22) | 106.46 (9.34) |
| Cognitive Status at last cognitive assessment, n(%) |  |  |  |  |
| CU-S | 283 (74.5) | 89 (81.7) | 57 (62.0) | 310 (70.9) |
| CU-D | 35 ( 9.2) | 20 (18.3) | 13 (14.1) | 45 (10.3) |
| Impaired_Not_MCI | 0 ( 0.0) | 0 ( 0.0) | 0 ( 0.0) | 0 ( 0.0) |
| MCI | 10 ( 2.6) | 0 ( 0.0) | 15 (16.3) | 23 ( 5.3) |
| Dementia | 0 ( 0.0) | 0 ( 0.0) | 5 ( 5.4) | 5 ( 1.1) |
| missing | 52 (13.7) | 0 ( 0.0) | 2 ( 2.2) | 54 (12.4) |
| *N's will vary by cognitive outcome. |  |  |  |  |
| Abbreviations: A=Amyloid (-=negative; +=positive); APOE=apolipoprotein E; CU=Cognitively Unimpaired (subtypes: S=Stable; D=Declining); MCI=mild cognitive impairment; WRAT=Wide Range Achievement Test |  |  |  |  |

Figure 5 depicts slopes and 95% CI’s by amyloid group for the three study partner outcomes based on the CDR and/or QDRS. None of the study partner outcomes’ slopes differ from 0 in the A- and subA+ groups, while the slopes of the QDRS – SB and the QDRS Total score (10 items) differ from 0 in the A+ group (see also the bottom section of Supplementary Table 2 for the depicted slope(se’s) and contributing sample sizes across study partner outcomes).

**Figure 5:**
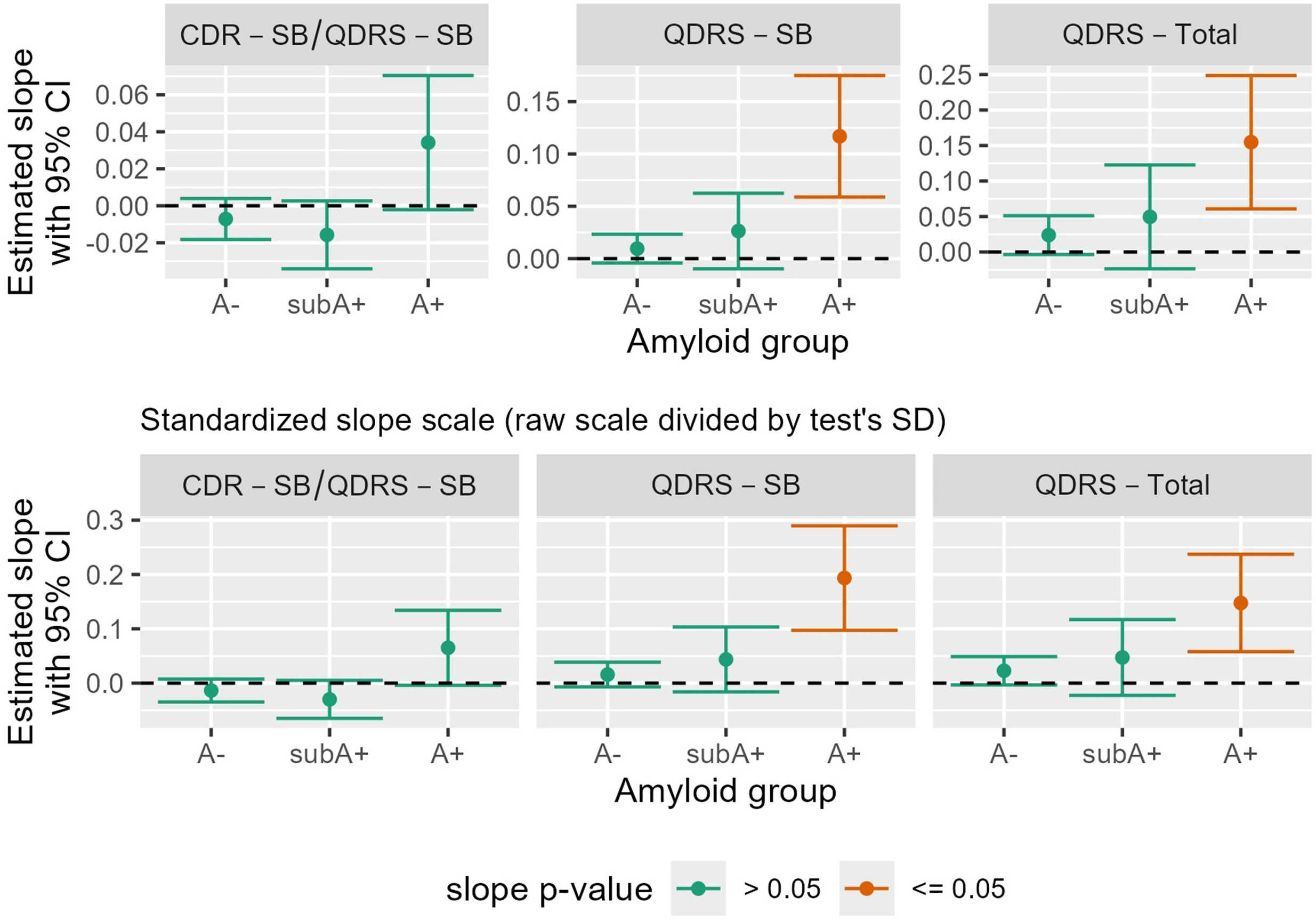
Study partner cognitive outcomes. Study partner (aka “informant”) reports of functioning (top row: average slopes and confidence intervals based on raw scores; bottom row: standardized slope scale to facilitate visual comparison of effect sizes across outcomes). CDR-SB/QDRS-SB is a sum of boxes score comprised of the CDR-SB when it is available or the sum of boxes on similar QDRS items when the CDR is not available (see Berman et al, 2017). Abbreviations: A- = amyloid negative group (CL<∼13); subA+ = sub-threshold to low positive group (CL ∼13-23); and A+ = overtly A+ group (CL >∼23); CL=Centiloid; CDR=Clinical Dementia Rating; CI= Confidence Interval; SB=Sum of Boxes; SD=Standard deviation; QDRS=Quick Dementia Rating Scale.

Sample size estimates to achieve 80% power to detect effect sizes ranging from 20-35% reduction in cognitive decline are summarized in Supplementary Table 5 for all cognitive outcomes (columns) and target groups (rows: subA+, attenuation to 0; subA+, attenuation to A-slope; A+ attenuation to 0; A+_<40CL_ attenuation to 0; and A+_>=40CL_ attenuation to 0). Given that many trials are often powered to detect a 25% reduction in decline, Figure 6 depicts sample size estimates for the cognitive outcomes that appear most efficient for each target treatment group: the top panel depicts the cognitive outcomes and sample size estimates for which <=2,000 people would be needed per treatment arm to detect a 25% slowing in cognitive decline in the subA+ group while the bottom panel depicts the same for the A+ group (and A+_<40CL_ and A+_>=40CL_ subsets). Figure 6 and Supplementary Table 5 illustrate that sample size needs vary widely depending on the cognitive outcome, the amyloid range targeted by the intervention and whether estimating attenuation towards 0 or towards the A-slope. In the SubA+ group, the outcomes requiring smallest sample sizes to detect a 25% reduction in slope included the Digit Symbol Substitution test (∼N=404/treatment arm) and the three composites (∼N of 1716-1893 per treatment arm). For the SubA+ group, targeting attenuation to the A-group slope instead of 0 slope increased sample sizes to 1656/per treatment group for Digit Symbol, 4786 for PACC3, 1783 for PACC5 (the least impacted by accounting for attenuation to the A-slope), and 2782 for PS, due to more similar slope behavior between the A- and SubA+ groups for these outcomes. For a trial targeting amyloid levels in the A+ group and corresponding subgroups, Digit Symbol, PACC3, and PACC5 again required the fewest participants per treatment arm regardless of whether the target range was A+, A+_<40CL_ or A+_>=40CL._ Sample sizes to detect 25% effects on study partner reports showed the QDRS-SB and QDRS Total scores required sample sizes similar to the composites for the A+ group and the A+_>=40CL_ subgroup.

**Figure 6:**
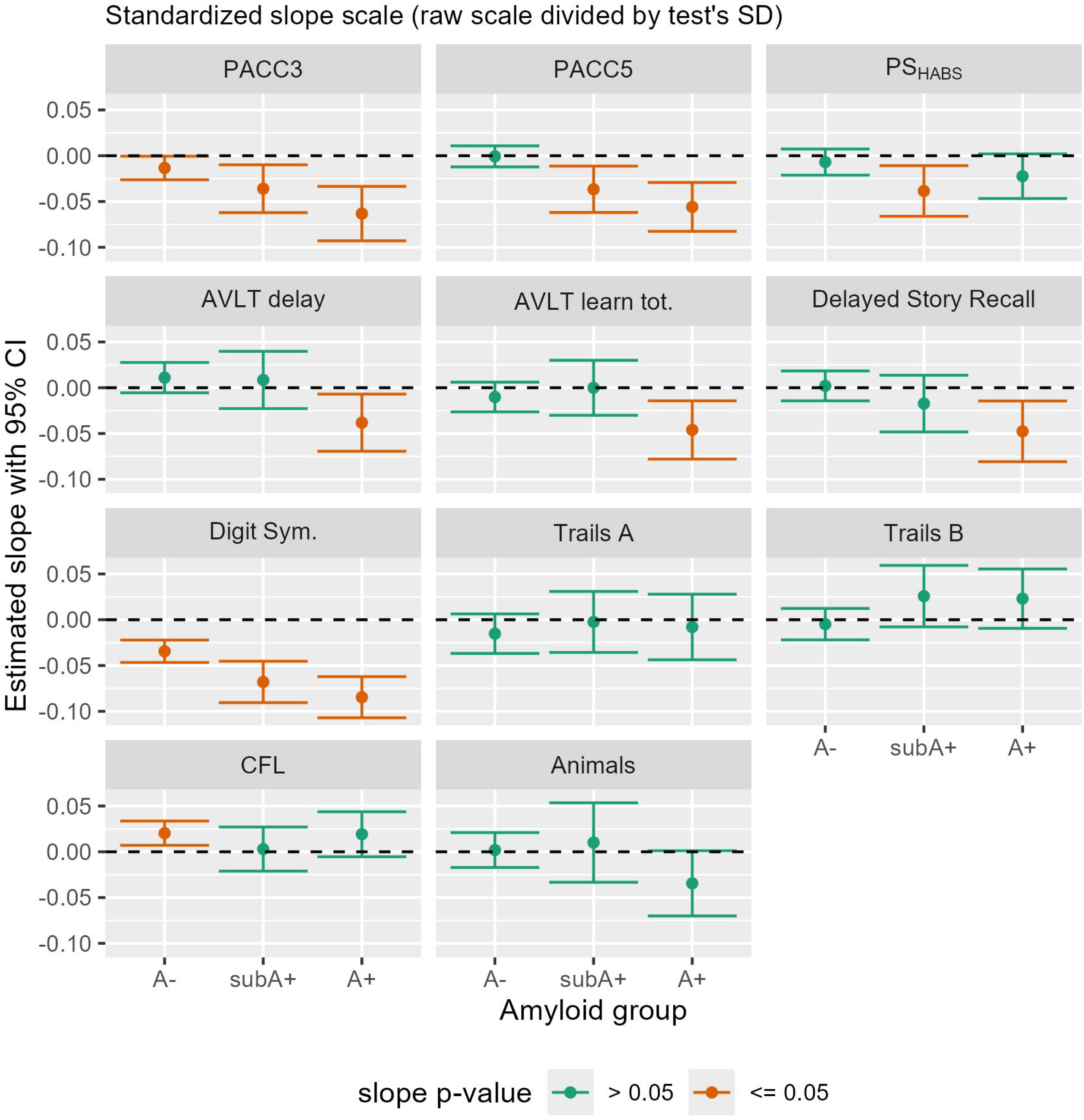
Sample size estimates for cognitive and study partner outcomes requiring <=2,000 per treatment arm. Top panel: Sample size estimates are shown for the subset of cognitive and study partner outcomes that required <=2,000 per treatment arm to detect a 25% slowing in decline in a subA+ study. When attenuation to the A-slope was factored in, only the Digit Symbol Substitution Task and PACC5 composite were sensitive enough to preclinical decline to require <=2000 per treatment arm. Bottom panel: Sample size estimates are shown for the subset of cognitive and study partner outcomes that required <=2,000 per treatment arm to detect a 25% slowing in decline in each of three A+ trial design options: Solid red line depicts sample sizes needed for 25% slowing in decline for all in the A+ group; Dotted lines with triangles or squares represent sample size estimates for the subsets that are <40 CL or >=40 CL, respectively. Sample size estimates for all cognitive and study partner outcomes and for a range of effect sizes are also listed in Supplementary Table 5. Abbreviations: A-= amyloid negative group (CL<∼13); subA+ = sub-threshold to low positive group (CL ∼13-23); and A+ = overtly A+ group (CL >∼23); CL = Centiloid; DVR=Distribution Volume Ratio; pTau217= phosphorylated tau protein at amino acid 217; AVLT: Rey Auditory Verbal Learning Test; Digit Sym: CFL: Phonemic fluency with letters C, F, and L; CI= Confidence Interval; Digit Sym.=Digit Symbol Substitution Test; Logical Mem=Logical Memory II; PACC: Preclinical Alzheimer’s Cognitive Composite (3 and 5 test versions); PS: Processing Speed composite (inspired by Harvard Aging Brain Study (HABS) composite).

In sensitivity analysis including a sex x age interaction in the estimated outcome trajectories of the amyloid groups, no significant differences were observed in trajectories between men and women for any outcome or amyloid subgroup.

## 4 Discussion

These analyses used the well-characterized Wisconsin Registry for Alzheimer’s Prevention (WRAP) study to obtain sample size estimates for preclinical amyloid clearance trials enrolling participants who are cognitively unimpaired at baseline and have either sub-threshold/low positive levels of amyloid (“subA+”) or more strongly positive (“A+”) baseline amyloid PET levels. To obtain sample size estimates, we first used longitudinal WRAP data to estimate change rates in amyloid PET, plasma p-tau217, and cognitive outcomes sensitive to preclinical or clinical change. We observed that change varied by outcome and PET amyloid group (A-, subA+, and A+) with least cognitive change in the A-group and greatest change in the A+ group across most outcomes. We then used these change estimates to obtain sample size estimates for each arm of a two-armed amyloid clearance trial (80% power). Very small samples would be needed to detect meaningful reductions in PET amyloid slopes and slightly to much larger sample sizes would be needed to detect changes in plasma p-tau217 slopes over a 3-year follow-up window. Larger sample sizes were generally needed to detect meaningful slowing of cognitive decline over a 6-year follow-up window ; sample size requirements varied widely by choice of cognitive outcome, the amyloid levels of the hypothetical treatment groups, and whether attenuation in decline was relative to zero or to “normal aging” rates estimated by the A-group. Sample size estimates based on study partner reports suggested such measures may also be useful in preclinical trials targeting the A+ range (but not trials targeting the subA+ range).

### 4.1 Slopes and sample size projections for amyloid biomarkers

WRAP PiB slope estimates were consistent with literature showing an inflection point of ∼12-13 CL.(52, 53) Specifically, while the slope in the A-subset did not differ from 0 and had limited variability, the slopes were positive in both the subA+ and A+ subsets and corresponded to approximately 4-5 CL increases per year (based on conversion formula in (21)); before adjusting for potential drop-outs, fewer than 50 per group would be needed to have power to detect 60-90% treatment effects in each of these groups. Results from trials of symptomatic participants (i.e., those with MCI or mild dementia) suggest that amyloid clearance is more likely to be achieved in those who begin with lower amyloid levels(57) while other studies have shown that higher levels of amyloid PET burden are associated with longer presence of amyloid (e.g., (21)), greater preclinical cognitive decline (e.g., (20)), and more tau abnormalities (e.g., (58)). Our study also showed increased tau abnormalities across the amyloid groups. Specifically, using a pTau217_ALZpath_ threshold of >0.64 pg/mL for tau PET positive,(10) the n(%) of T+ across the amyloid groups was 4(1.7%) for A-, 4(9.8%) for subA+ and 32(50.0%) for A+. Several studies including WRAP(16, 58) have shown faster preclinical cognitive decline in those with both elevated amyloid and tau neuropathology. Thus, intervening at the earliest stages of detectable amyloid PET signal may be most fruitful for delaying or preventing AD dementia.

The positive plasma p-tau217 slopes in the A+ group were consistent with the growing literature showing strong associations between p-tau217 and amyloid PET positivity (e.g., (10, 24, 25)). Estimates indicated that both assays would require very reasonable sample sizes (<100 per treatment group before adjusting for dropouts) to detect 60-90% improvements in trajectories in those at or above the A+ threshold used here (i.e., >=23 CL). Improvements in plasma p-tau217 trajectories have been reported in amyloid clearing trials for more advanced stages. For example, Pontecorvo et al (12) reported p-tau217 improvements as early as 12 weeks after beginning treatment with donanemab in the TRAILBLAZER-ALZ trial; these differences correlated with amyloid PET improvements as well, suggesting that plasma p-tau217 may be a viable measure of treatment success. Across all three biomarkers, sample size estimates were <100 per treatment arm for both the A+_<40 CL_ and A+_>=40 CL_ subgroups.

Slope estimates and sample size projections for the two plasma p-tau217 assays suggest different things. The positive pTau217_Meso_ slopes in the subA+ group were associated with projected sample size needs of ∼200 per treatment arm if effect size estimates are relative to attenuation to 0 and ∼90 per arm if effect size estimates are relative to attenuation to the average pTau217_Meso_ A-slope. In contrast, sample size estimates for pTau217_ALZpath_ are >800 per arm to detect 60-90% attenuation to 0, and the slope estimate in the subA+ group are lower than in the A-group. Together, these results suggest that the ALZpath plasma p-tau 217 assay may be less viable as an outcome for trials addressing the earliest signs of amyloid PET burden. As noted by others (e.g., (59)) additional studies are needed to understand how various assays perform in diverse samples.

### 4.2 Slopes and sample size projections for cognitive outcomes

Cognitive slope estimates showed that even among cognitively unimpaired persons, presumably AD-related declines are already detectable over a six-year window for Digit Symbol Substitution and PACC scores in the subA+ subset with significant preclinical declines emerging next in immediate learning and delayed recall in the A+ group. The Digit Symbol Substitution test, a measure of executive function, attention, processing speed and other domains(60), has been shown to have good sensitivity to cognitive dysfunction and risk of dementia.(60)

In both trial scenarios (i.e., CU, subA+ participants or CU, A+ participants), Digit Symbol was associated with the smallest projected sample size needs (e.g., for a 25% reduction in decline, 404 per subA+ treatment arm or 264 per A+ treatment arm). Given that Digit Symbol slopes were negative in the A-group, sample size projections were approximately quadrupled when effect sizes were calculated relative to A-slopes (n=1656 vs 404). Not factoring in such “normal” rates of change may contribute to negative trials related to lower than projected statistical power.

The next most efficient cognitive outcome for both trial designs was PACC5. This composite, includes the three tests of the WRAP PACC3 (AVLT sum of learning trials, Logical Memory II, and Digit Symbol Substitution Test) plus phonemic fluency (CFL) and MMSE, two measures that showed minimal age-related change in an early analysis of unimpaired WRAP participants(26). The PACC5 slope in the A-subset did not differ from 0 and sample size projects for the subA+ group were similar whether the effect size was based on attenuation to A-(n=1783 per arm) or simply reduced decline relative to the subA+ observed slope (n=1716). Thus, it is important to understand how a test or composite performs in comparable biomarker negative groups. In the A+ group, sample size projections were 838 per arm for anyone in the A+ group, with 1079 for a subgroup <40 CL at baseline and 507 for those with CL==40 at baseline. The WRAP PACC5 components are quite similar to the ones used in the PACC5 for the AHEAD 3-45 studies.(7, 61) The AHEAD 3 cohort closely resembles the <40 CL subset of our A+ group; for that study, the primary outcome was amyloid PET and 400 participants were enrolled for four years of follow-up. The authors mention that cognitive decline would be too slow to make it a viable outcome in a four-year study. Our projections were based on a six year window per recommendations by Insel et al.(5)

None of the study partner measures’ slopes differed from zero in the A- or subA+ groups; the largest slopes were observed in the A+ group and for the sum of the first six QDRS items (i.e., a score approximate to the CDR-SB) followed by the QDRS total score. Similar patterns were also reported by Duff et al (62) in an older sample (average baseline age of 74 years). WRAP-based sample size estimates and the ease of QDRS administration may make this attractive to include in a CU, A+ clinical trial.

### 4.3 Sensitivity analyses

While no significant trajectory differences were observed in the data for male vs female subjects, as with any null finding, it is possible that we were not adequately powered to detect an interaction with our data. Analyses by Insel et al(5) identified a sex by amyloid interaction in association with PACC trajectories in one of three cohorts they examined; all three cohorts they examined were about a decade older on average at baseline than the WRAP sample.

### 4.4 Strengths, Limitations, and Conclusions

WRAP’s rich longitudinal biomarker and cognitive data provide unique opportunity to characterize cognitive change across multiple outcomes by amyloid PET level in people who are cognitively unimpaired at cognitive baseline. In the CU/A-subset, several cognitive outcomes showed small but statistically significant rates of non-AD decline. Sample size projections demonstrate how not accounting for this rate of decline may underestimate sample size needs. Limitations include small cell sizes for some slope estimates, a predominantly non-Hispanic White sample, and a simple approach to estimating sample sizes (i.e., t-test with no adjustments for drop-outs). While sample size estimates may vary with a different analysis method and will increase when adjusted for assumed drop-out rates, the relative comparisons (i.e., across outcomes or trial scenarios) are useful and illustrate that a relatively simple cognitive battery could be used to obtain cognitive outcomes that are well-suited to detecting slowing in preclinical change over a six-year period; powering a study to detect 25% slowing in cognitive decline would more than adequately power larger treatment effects in plasma or PET markers of amyloid.

## Data Availability

All data produced in the present study are available upon reasonable request to the authors. In addition, information about the data requestion process for Wisconsin Registry for Alzheimer's Prevention data is found here: https://wrap.wisc.edu/data-requests-2/

## Acknowledgements

The authors wish to thank the WRAP study participants and their study partners for their contributions to the study; without their dedication, none of this research would be possible. The authors also wish to thank the broader WRAP team (including program managers, study coordinators, lab technicians, and all others involved in making the study happen), the WADRC blood biomarker team, and the WRAP and WADRC data and imaging teams.

## Consent Statement

Participants provided informed consent for all aspects of data collection, and all study procedures were conducted in accordance with the Helsinki Declaration and approved by the University of Wisconsin Institutional Review Board. **Key Words:** clinical trial design, preclinical intervention, Alzheimer’s disease, biomarkers, preclinical

## References

1. Birdsill AC, Koscik RL, Cody KA, Jonaitis EM, Cadman RV, Erickson CM, et al. Trajectory of clinical symptoms in relation to amyloid chronicity. Alzheimers Dement (Amst). 2022;14(1):e12360.

2. Caselli RJ, Langlais BT, Dueck AC, Chen Y, Su Y, Locke DEC, et al. Neuropsychological decline up to 20 years before incident mild cognitive impairment. Alzheimer’s & Dementia. 2019;1552–5260.

3. Payton NM, Marseglia A, Grande G, Fratiglioni L, Kivipelto M, Bäckman L, et al. Trajectories of cognitive decline and dementia development: A 12-year longitudinal study. Alzheimer’s & Dementia. 2023;19(3):857–67 1552-5260.

4. Donohue MC, Sperling RA, Salmon DP, Rentz DM, Raman R, Thomas RG, et al. The Preclinical Alzheimer Cognitive Composite: Measuring Amyloid-Related Decline. JAMA Neurology. 2014;71(8):961–70 2168-6149.

5. Insel PS, Weiner M, Mackin RS, Mormino E, Lim YY, Stomrud E, et al. Determining clinically meaningful decline in preclinical Alzheimer disease. Neurology. 2019;93(4):e322–e33 0028-3878.

6. Budd Haeberlein S, Aisen PS, Barkhof F, Chalkias S, Chen T, Cohen S, et al. Two randomized phase 3 studies of aducanumab in early Alzheimer’s disease. The journal of prevention of Alzheimer’s disease. 2022;9(2):197–210 2426-0266.

7. Rafii MS, Sperling RA, Donohue MC, Zhou J, Roberts C, Irizarry MC, et al. The AHEAD 3-45 Study: design of a prevention trial for Alzheimer’s disease. Alzheimer’s & dementia. 2023;19(4):1227–33 552-5260.

8. Jutten RJ, Papp KV, Hendrix S, Ellison N, Langbaum JB, Donohue MC, et al. Why a clinical trial is as good as its outcome measure: A framework for the selection and use of cognitive outcome measures for clinical trials of Alzheimer’s disease. Alzheimer’s & Dementia. 2023;19(2):708–20 1552-5260.

9. Jonaitis EM, Janelidze S, Cody KA, Langhough R, Du L, Chin NA, et al. Plasma phosphorylated tau 217 in preclinical Alzheimer’s disease. Brain Commun. 2023;5(2):fcad057.

10. Ashton NJ, Brum WS, Di Molfetta G, Benedet AL, Arslan B, Jonaitis E, et al. Diagnostic Accuracy of a Plasma Phosphorylated Tau 217 Immunoassay for Alzheimer Disease Pathology. JAMA Neurol. 2024;81(3):255–63.

11. Devanarayan V, Charil A, Horie K, Doherty T, Llano DA, Andreozzi E, et al. Plasma pTau217 ratio predicts continuous regional brain tau accumulation in amyloid-positive early Alzheimer’s disease. Alzheimer’s & Dementia. 2025;21(2):e14411.

12. Pontecorvo MJ, Lu M, Burnham SC, Schade AE, Dage JL, Shcherbinin S, et al. Association of donanemab treatment with exploratory plasma biomarkers in early symptomatic Alzheimer disease: a secondary analysis of the TRAILBLAZER-ALZ randomized clinical trial. JAMA neurology. 2022;79(12):1250–9 2168-6149.

13. Ashton NJ, Janelidze S, Mattsson-Carlgren N, Binette AP, Strandberg O, Brum WS, et al. Differential roles of Abeta42/40, p-tau231 and p-tau217 for Alzheimer’s trial selection and disease monitoring. Nat Med. 2022;28(12):2555–62.

14. Sperling RA, Donohue MC, Rissman RA, Johnson KA, Rentz DM, Grill JD, et al. Amyloid and Tau Prediction of Cognitive and Functional Decline in Unimpaired Older Individuals: Longitudinal Data from the A4 and LEARN Studies. The Journal of Prevention of Alzheimer’s Disease. 2024;11(4):802–13.

15. Johnson SC, Koscik RL, Jonaitis EM, Clark LR, Mueller KD, Berman SE, et al. The Wisconsin Registry for Alzheimer’s Prevention: A review of findings and current directions. Alzheimers Dement (Amst). 2018;10:130–42.

16. Betthauser TJ, Koscik RL, Jonaitis EM, Allison SL, Cody KA, Erickson CM, et al. Amyloid and tau imaging biomarkers explain cognitive decline from late middle-age. Brain. 2020;143(1):320–35.

17. Clark LR, Racine AM, Koscik RL, Okonkwo OC, Engelman CD, Carlsson CM, et al. Beta-amyloid and cognitive decline in late middle age: Findings from the Wisconsin Registry for Alzheimer’s Prevention study. Alzheimers Dement. 2016;12(7):805–14.

18. Johnson SC, Christian BT, Okonkwo OC, Oh JM, Harding S, Xu G, et al. Amyloid burden and neural function in people at risk for Alzheimer’s Disease. Neurobiology of aging. 2014;35(3):576–84.

19. Racine AM, Clark LR, Berman SE, Koscik RL, Mueller KD, Norton D, et al. Associations between Performance on an Abbreviated CogState Battery, Other Measures of Cognitive Function, and Biomarkers in People at Risk for Alzheimer’s Disease. J Alzheimers Dis. 2016;54(4):1395–408.

20. Koscik RL, Betthauser TJ, Jonaitis EM, Allison SL, Clark LR, Hermann BP, et al. Amyloid duration is associated with preclinical cognitive decline and tau PET. Alzheimers Dement (Amst). 2020;12(1):e12007.

21. Betthauser TJ, Bilgel M, Koscik RL, Jedynak BM, An Y, Kellett KA, et al. Multi-method investigation of factors influencing amyloid onset and impairment in three cohorts. Brain. 2022;145(11):4065–79.

22. Schindler SE, Li Y, Buckles VD, Gordon BA, Benzinger TLS, Wang G, et al. Predicting symptom onset in sporadic Alzheimer disease with amyloid PET. Neurology. 2021;97(18):e1823–e34 0028-3878.

23. Therneau TM, Knopman DS, Lowe VJ, Botha H, Graff-Radford J, Jones DT, et al. Relationships between β-amyloid and tau in an elderly population: An accelerated failure time model. Neuroimage. 2021;242:118440 1053–8119.

24. Jonaitis EM, Janelidze S, Cody KA, Langhough R, Du L, Chin NA, et al. Plasma phosphorylated tau 217 in preclinical Alzheimer’s disease. Brain Communications. 2023;5(2 2632-1297).

25. Du L, Langhough RE, Wilson RE, Reyes RER, Hermann BP, Jonaitis EM, et al. Longitudinal plasma phosphorylated-tau217 and other related biomarkers in a non-demented Alzheimer’s risk-enhanced sample. Alzheimers Dement. 2024;20(9):6183–204.

26. Jonaitis EM, Koscik RL, Clark LR, Ma Y, Betthauser TJ, Berman SE, et al. Measuring longitudinal cognition: Individual tests versus composites. Alzheimers Dement (Amst). 2019;11:74–84.

27. Koscik RL, Norton DL, Allison SL, Jonaitis EM, Clark LR, Mueller KD, et al. Characterizing the Effects of Sex, APOE varepsilon4, and Literacy on Mid-life Cognitive Trajectories: Application of Information-Theoretic Model Averaging and Multi-model Inference Techniques to the Wisconsin Registry for Alzheimer’s Prevention Study. J Int Neuropsychol Soc. 2019;25(2):119–33.

28. Jonaitis EM, Hermann BP, Mueller KD, Clark LR, Du L, Betthauser TJ, et al. Longitudinal normative standards for cognitive tests and composites using harmonized data from two Wisconsin AD-risk-enriched cohorts. Alzheimers Dement. 2024;20(5):3305–21.

29. Neu SC, Pa J, Kukull W, Beekly D, Kuzma A, Gangadharan P, et al. Apolipoprotein E Genotype and Sex Risk Factors for Alzheimer Disease: A Meta-analysis. JAMA Neurol. 2017;74(10):1178–89.

30. Schmidt M. Rey auditory verbal learning test: Western Psychological Services Los Angeles; 1996.

31. Wechsler D. WAIS-R: Manual: Wechsler adult intelligence scale-revised. (No Title). 1981.

32. Reitan RM. Validity of the Trail Making Test as an indicator of organic brain damage. Perceptual and motor skills. 1958;8(3):271–6 0031-5125.

33. Wechsler D. Wechsler memory scale-revised. Psychological Corporation. 1987.

34. Bruno D, Mueller KD, Betthauser T, Chin N, Engelman CD, Christian B, et al. Serial position effects in the Logical Memory Test: Loss of primacy predicts amyloid positivity. J Neuropsychol. 2021;15(3):448–61.

35. Rosen WG. Verbal fluency in aging and dementia. Journal of clinical and experimental neuropsychology. 1980;2(2):135–46 0165-475.

36. Farrell ME, Papp KV, Buckley RF, Jacobs HIL, Schultz AP, Properzi MJ, et al. Association of emerging β-amyloid and tau pathology with early cognitive changes in clinically normal older adults. Neurology. 2022;98(15):e1512–e24 0028-3878.

37. Morris JC. Clinical dementia rating: a reliable and valid diagnostic and staging measure for dementia of the Alzheimer type. International psychogeriatrics. 1997;9(S1):173–6 1741-203X.

38. O’Bryant SE, Waring SC, Cullum CM, Hall J, Lacritz L, Massman PJ, et al. Staging dementia using Clinical Dementia Rating Scale Sum of Boxes scores: a Texas Alzheimer’s research consortium study. Archives of neurology. 2008;65(8):1091–5 0003-9942.

39. Galvin JE. The Quick Dementia Rating System (QDRS): a rapid dementia staging tool. Alzheimer’s & Dementia: Diagnosis, Assessment & Disease Monitoring. 2015;1(2):249–59 2352-8729.

40. Berman SE, Koscik RL, Clark LR, Mueller KD, Bluder L, Galvin JE, et al. Use of the Quick Dementia Rating System (QDRS) as an Initial Screening Measure in a Longitudinal Cohort at Risk for Alzheimer’s Disease. J Alzheimers Dis Rep. 2017;1(1):9–13.

41. Huang Q, Bolt D, Jonaitis E, Hermann B, Studer R, Du L, et al. Performance of study partner reports in a non-demented at-risk sample. Alzheimers Dement. 2024.

42. Langhough Koscik R, Hermann BP, Allison S, Clark LR, Jonaitis EM, Mueller KD, et al. Validity evidence for the research category,“cognitively unimpaired–declining,” as a risk marker for mild cognitive impairment and Alzheimer’s disease. Frontiers in Aging Neuroscience. 2021;13:688478 1663–4365.

43. Albert MS, DeKosky ST, Dickson D, Dubois B, Feldman HH, Fox NC, et al. The diagnosis of mild cognitive impairment due to Alzheimer’s disease: Recommendations from the National Institute on Aging-Alzheimer’s Association workgroups on diagnostic guidelines for Alzheimer’s disease. Alzheimer’s & Dementia. 2011;7(3 1552-5260):270-9.

44. McKhann GM, Knopman DS, Chertkow H, Hyman BT, Jack Jr CR, Kawas CH, et al. The diagnosis of dementia due to Alzheimer’s disease: Recommendations from the National Institute on Aging-Alzheimer’s Association workgroups on diagnostic guidelines for Alzheimer’s disease. Alzheimer’s & dementia. 2011;7(3):263–9 1552-5260.

45. Sprecher KE, Bendlin BB, Racine AM, Okonkwo OC, Christian BT, Koscik RL, et al. Amyloid burden is associated with self-reported sleep in nondemented late middle-aged adults. Neurobiol Aging. 2015;36(9):2568–76.

46. Klunk WE, Koeppe RA, Price JC, Benzinger TL, Devous Sr MD, Jagust WJ, et al. The Centiloid Project: standardizing quantitative amyloid plaque estimation by PET. Alzheimer’s & dementia. 2015;11(1):1–15. e4 1552-5260.

47. Zhang H, Liu J, Zhang N, Jeromin A, Lin ZJ. Validation of an Ultra-Sensitive Method for Quantitation of Phospho-Tau 217 (pTau217) in Human Plasma, Serum, and CSF using the ALZpath pTau217 Assay on the Quanterix HD-X Platform. The Journal of Prevention of Alzheimer’s Disease. 2024;11(5):1206–11.

48. R Development Core Team. R: A language and environment for statistical computing. Vienna, Austria: R Foundation for Statistical Computing; 2010. p. Retrieved from http://www.R-project.org.

49. Bates D, Mächler M, Bolker B, Walker S. Fitting linear mixed-effects models using lme4. Journal of statistical software. 2015;67:1–48.

50. Yoshida K, Bartel A, Chipman J. tableone: create “Table 1” to describe baseline characteristics with or without propensity score weights. R package version 013. 2022;2.

51. Wickham H. Getting Started with ggplot2. ggplot2: Elegant graphics for data analysis: Springer; 2016. p. 11–31.

52. Salvadó G, Molinuevo JL, Brugulat-Serrat A, Falcon C, Grau-Rivera O, Suárez-Calvet M, et al. Centiloid cut-off values for optimal agreement between PET and CSF core AD biomarkers. Alzheimer’s research & therapy. 2019;11:1–12.

53. Burnham SC, Cox T, Benzinger T, Bourgeat P, Cruchaga C, Doecke JD, et al. When Does Alzheimer’s Disease Start? Robust Estimates Based on Longitudinal Aβ-Amyloid-PET in Three Large International Cohorts.

54. Hanseeuw BJ, Malotaux V, Dricot L, Quenon L, Sznajer Y, Cerman J, et al. Defining a Centiloid scale threshold predicting long-term progression to dementia in patients attending the memory clinic: an [18F] flutemetamol amyloid PET study. European Journal of Nuclear Medicine and Molecular Imaging. 2021;48(1):302–10.

55. Amadoru S, Doré V, McLean CA, Hinton F, Shepherd CE, Halliday GM, et al. Comparison of amyloid PET measured in Centiloid units with neuropathological findings in Alzheimer’s disease. Alzheimer’s research & therapy. 2020;12(1):22.

56. Farrell ME, Jiang S, Schultz AP, Properzi MJ, Price JC, Becker JA, et al. Defining the lowest threshold for amyloid-PET to predict future cognitive decline and amyloid accumulation. Neurology. 2021;96(4):e619–e31 0028-3878.

57. Shcherbinin S, Evans CD, Lu M, Andersen SW, Pontecorvo MJ, Willis BA, et al. Association of amyloid reduction after donanemab treatment with tau pathology and clinical outcomes: the TRAILBLAZER-ALZ randomized clinical trial. JAMA neurology. 2022;79(10):1015–24.

58. Cody KA, Langhough RE, Zammit MD, Clark L, Chin N, Christian BT, et al. Characterizing brain tau and cognitive decline along the amyloid timeline in Alzheimer’s disease. Brain. 2024;147(6):2144–57.

59. Teunissen CE, Kolster R, Triana-Baltzer G, Janelidze S, Zetterberg H, Kolb HC. Plasma p-tau immunoassays in clinical research for Alzheimer’s disease. Alzheimer’s & Dementia. 2025;21(1):e14397.

60. Jaeger J. Digit symbol substitution test: the case for sensitivity over specificity in neuropsychological testing. Journal of clinical psychopharmacology. 2018;38(5):513–9.

61. Papp KV, Rentz DM, Orlovsky I, Sperling RA, Mormino EC. Optimizing the preclinical Alzheimer’s cognitive composite with semantic processing: the PACC5. Alzheimer’s & Dementia: Translational Research & Clinical Interventions. 2017;3(4):668–77.

62. Duff K, Wan L, Embree L, Hoffman JM. Change in the Quick Dementia Rating System across time in older adults with and without cognitive impairment. Journal of Alzheimer’s Disease. 2023;93(2):449–57.

