## Supplementary material for "Use of preclinical Alzheimer’s disease trajectories for clinical trial design": Supplementary Materials.docx

**Supplementary Figure 1: Participant inclusion/exclusion criteria**

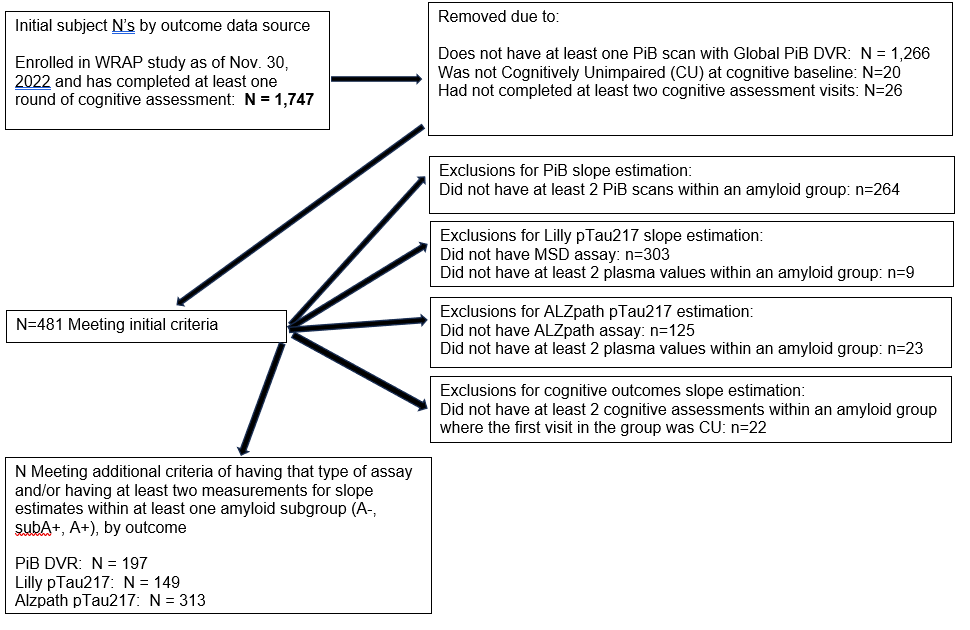

**Supplementary Figure 1 Note:** data pulled for these analyses consists primarily of the November 2022 freeze with two exceptions. 1) Plasma data were pulled on 1/23/2024 (from Panda) and 2) the CDR and QDRS data pull was updated on 12/4/2024 since these measures were added later to the study and benefitted from more data to inform the estimates.

**Supplementary Figure 2: Spaghetti plots of observed biomarkers vs age**

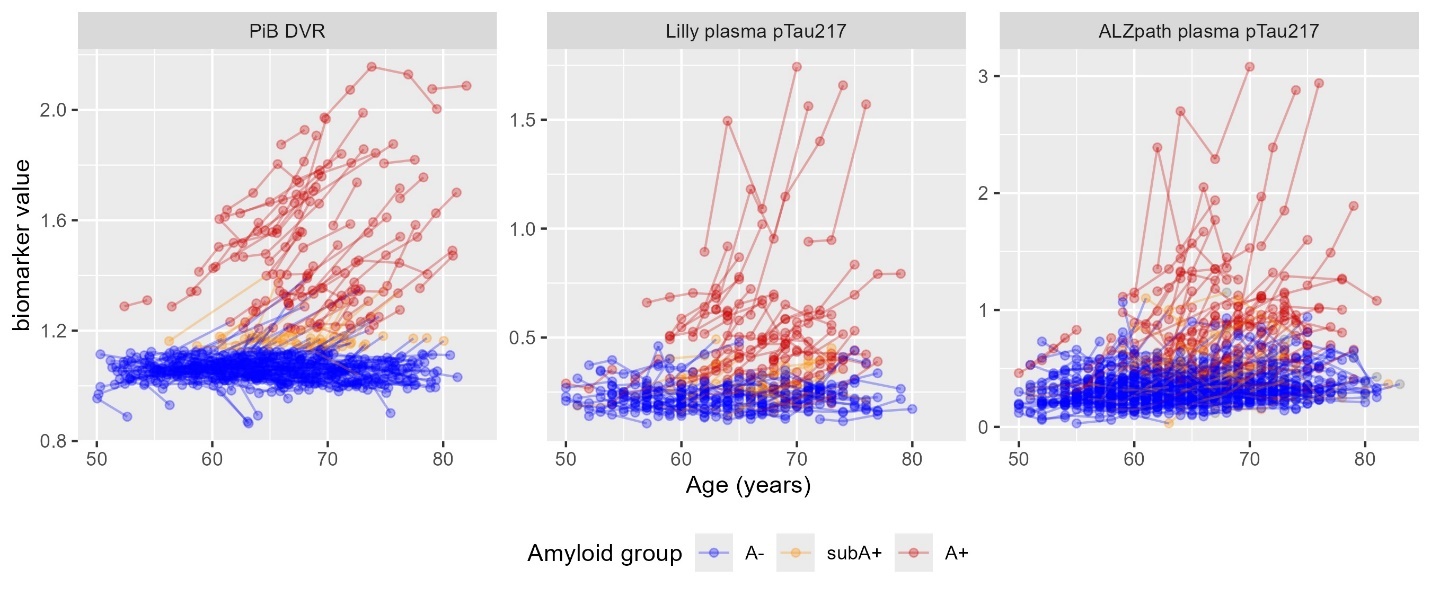

**Supplementary Figure 2 Note:** Spaghetti plots of observed biomarker data vs age: PiB (left), plasma pTau217_Meso_ (middle), and plasma pTau217_ALZpath_ (right). Blue, yellow, and red indicate A-, subA+, and A+, respectively. Abbreviations: A- = amyloid negative group (CL<~13); subA+ = sub-threshold to low positive group (CL ~13-23); and A+ = overtly A+ group (CL >~23); CL=Centiloid; DVR=Distribution Volume Ratio; PiB= Pittsburgh Compound B; pTau217= phosphorylated tau protein at amino acid 217; Lily MSD = Lily Meso Scale Discovery platform.

**Supplementary Figure 3: Spaghetti plots of observed plasma p-tau217 vs SILA model-estimated PiB DVR at plasma measurement.**

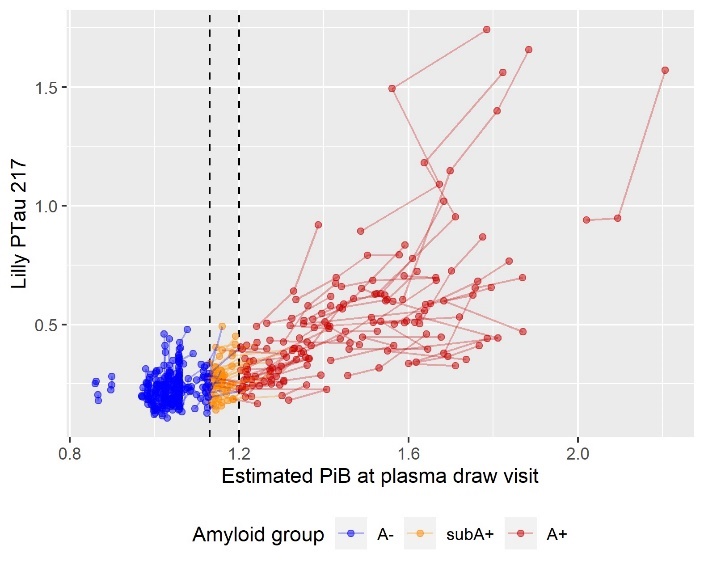

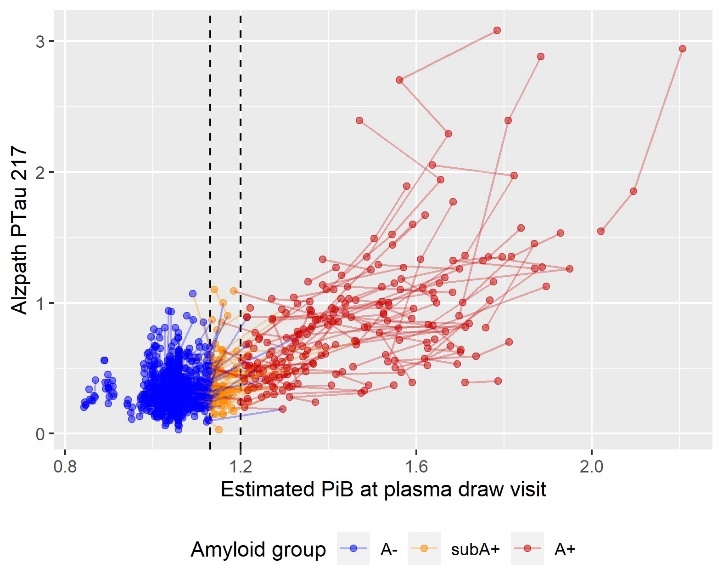

**Supplementary Figure 3 Note:** Spaghetti plots of observed plasma p-tau217 vs model-estimated PiB DVR at plasma measurement: Lily pTau217 (left) and ALZpath pTau217 (right) pg/mL. Abbreviations: A- = amyloid negative group (CL<~13); subA+ = sub-threshold to low positive group (CL ~13-23); and A+ = overtly A+ group (CL >~23); DVR=Distribution Volume Ratio; PiB= Pittsburgh Compound B; pTau217= phosphorylated tau protein at amino acid 217; Lily MSD = Lily Meso Scale Discovery platform; SILA=sampled iterative local approximation.

**Supplementary Figure 4: Average slope (and 95% Confidence Interval) by amyloid group**

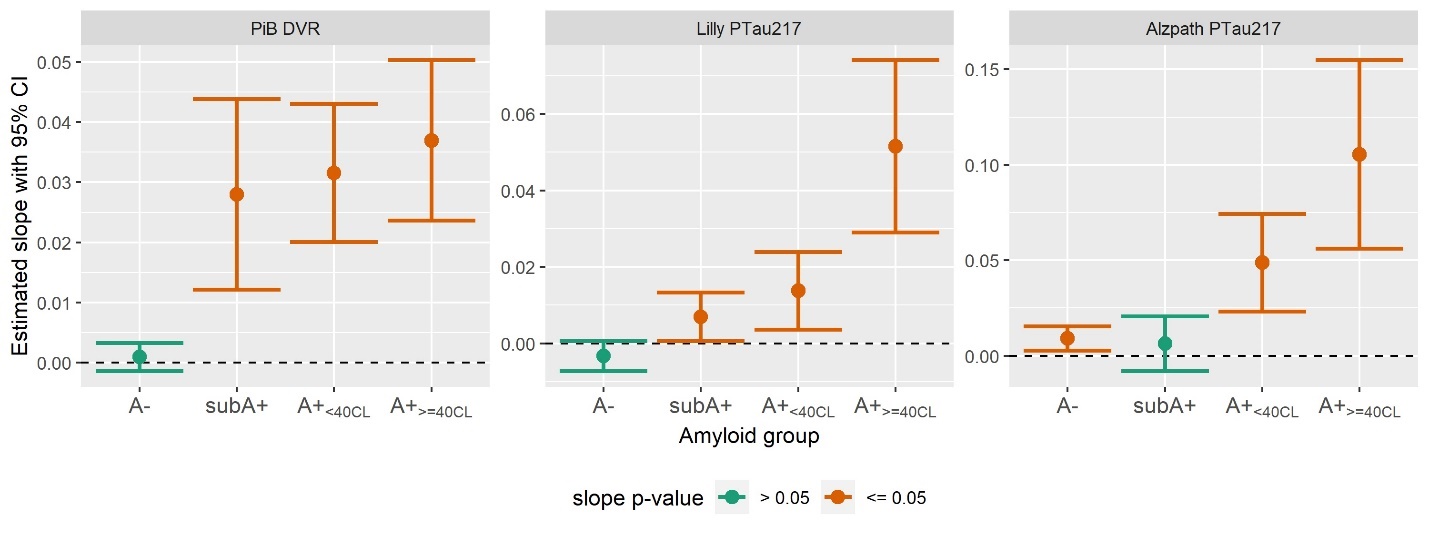

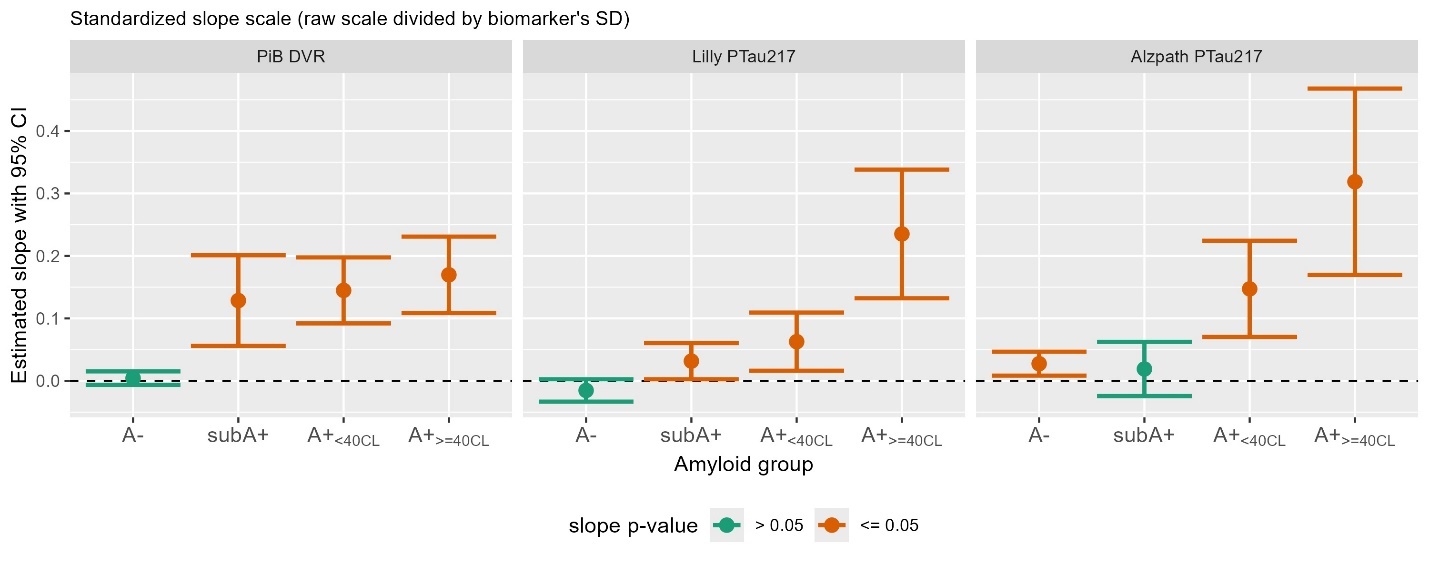

**Supplementary Figure 4 Note:** The top row depicts estimated slopes by amyloid group for PiB DVR (left), Lilly MSD plasma pTau217 pg/mL (middle), and Quanterix ALZpath plasma pTau217 pg/mL (right). The bottom row shows the same data using standardized slope estimates to facilitate visual comparisons of effect sizes across biomarkers. Slopes that differ significantly from 0 are in orange while those not differing from 0 are in green. This figure extends Figure 3 from the main manuscript by subdividing the A+ group into those who are <40CL or >=40 CL. Abbreviations: A- = amyloid negative group (CL<~13); subA+ = sub-threshold to low positive group (CL ~13-23); and A+ = overtly A+ group (CL >~23); CL=Centiloid; DVR=Distribution Volume Ratio; MSD= Meso Scale Discovery; PiB= Pittsburgh Compound B; pTau217= phosphorylated tau protein at amino acid 217.

**Supplementary Figure 5: Main manuscript Figure 3 revisited in the subA+ subset**

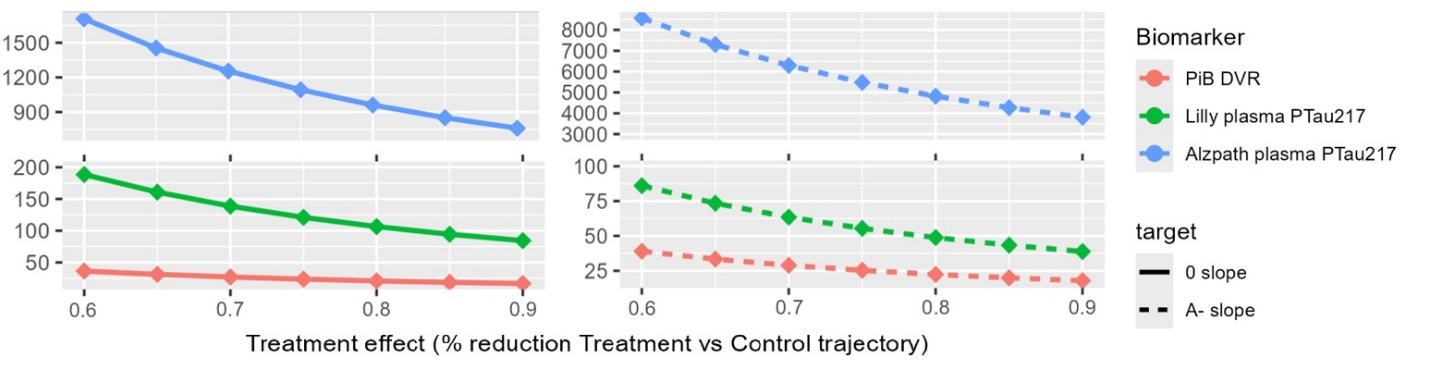

**Supplementary Figure 5 Note:** Given the large estimates for ALZpath pTau217 in this amyloid group, this figure reproduces main manuscript Figure 3 with breaks in the y-axis to allow closer inspection of sample size estimates for PiB DVR and pTau217_Meso_. Abbreviations: subA+ = sub-threshold to low positive group (CL ~13-23); CL=Centiloid; DVR=Distribution Volume Ratio; Lily MSD= Meso Scale Discovery; PiB= Pittsburgh Compound B; pTau217= phosphorylated tau protein at amino acid 217.

**Supplementary Figure 6: Standardized slope estimates for cognitive outcomes.**

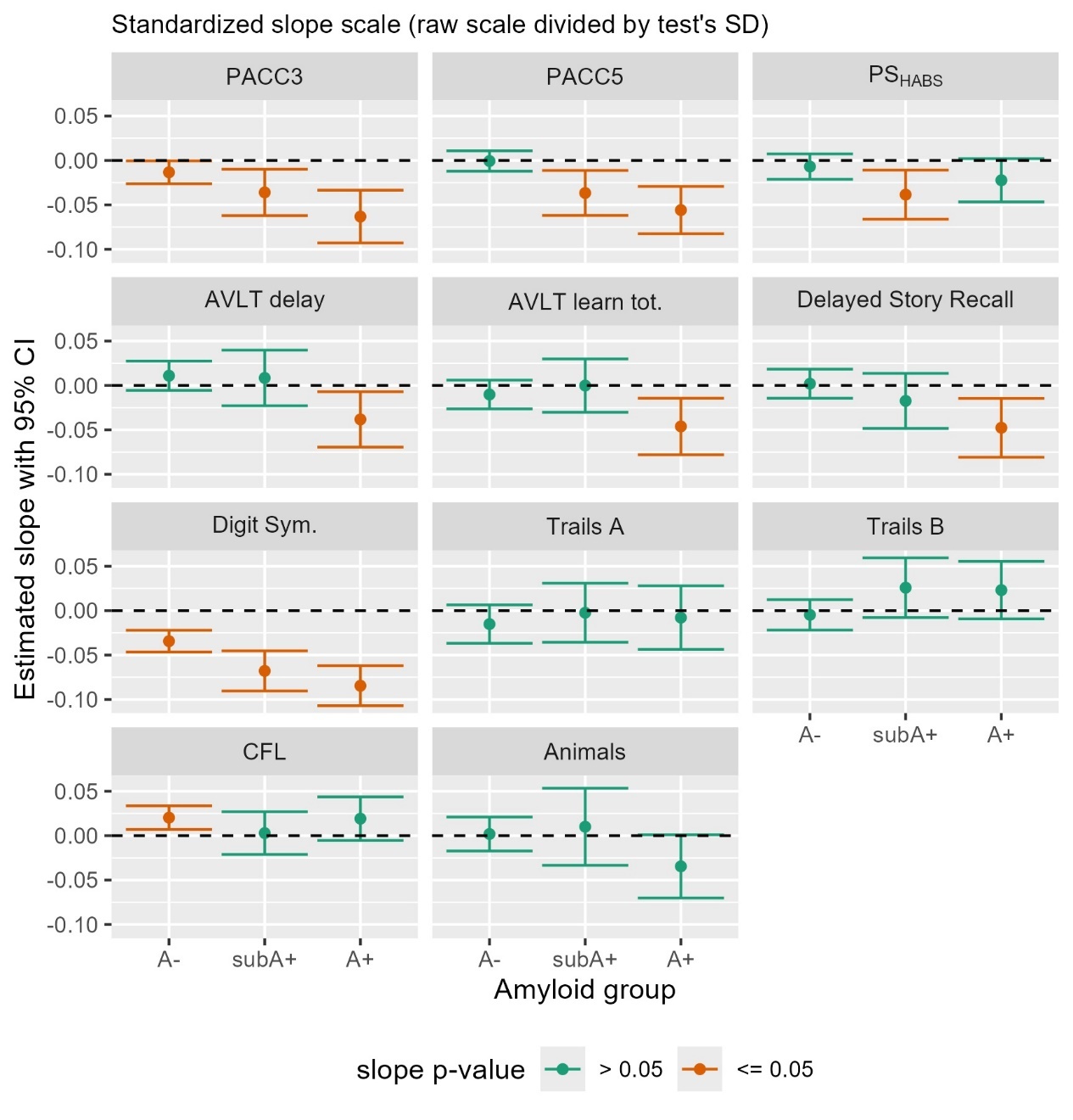

**Supplementary Figure 6 Note:** This figure is parallel to Figure 4 in the main manuscript. In Figure 4, the observed slopes and 95% confidence intervals (CIs) were shown using raw values from each test. Here, those slopes and CI’s have been converted to a standardized scale to allow visual comparison of effect sizes across cognitive outcomes. As in Figure 4, the top row depicts the cognitive composites and the bottom three rows show individual tests, by amyloid group. Green indicates slope did not differ from 0 (p>0.05) while orange indicates slopes that differed from 0. For all but Trails A and B, negative numbers indicate decline. Abbreviations: A- = amyloid negative group (CL<~13); subA+ = sub-threshold to low positive group (CL ~13-23); and A+ = overtly A+ group (CL >~23); CL=Centiloid; Animals: Semantic fluency with animal naming; AVLT: Rey Auditory Verbal Learning Test; Digit Sym: CFL: Phonemic fluency with letters C, F, and L; CI= Confidence Interval; Digit Sym.=Digit Symbol Substitution Test; PACC: Preclinical Alzheimer’s Cognitive Composite (3 and 5 test versions); PS: Processing Speed composite (inspired by Harvard Aging Brain Study (HABS) composite).

**Supplementary Figure 7: Supplementary Figure 4 revisited now depicting standardized slopes**

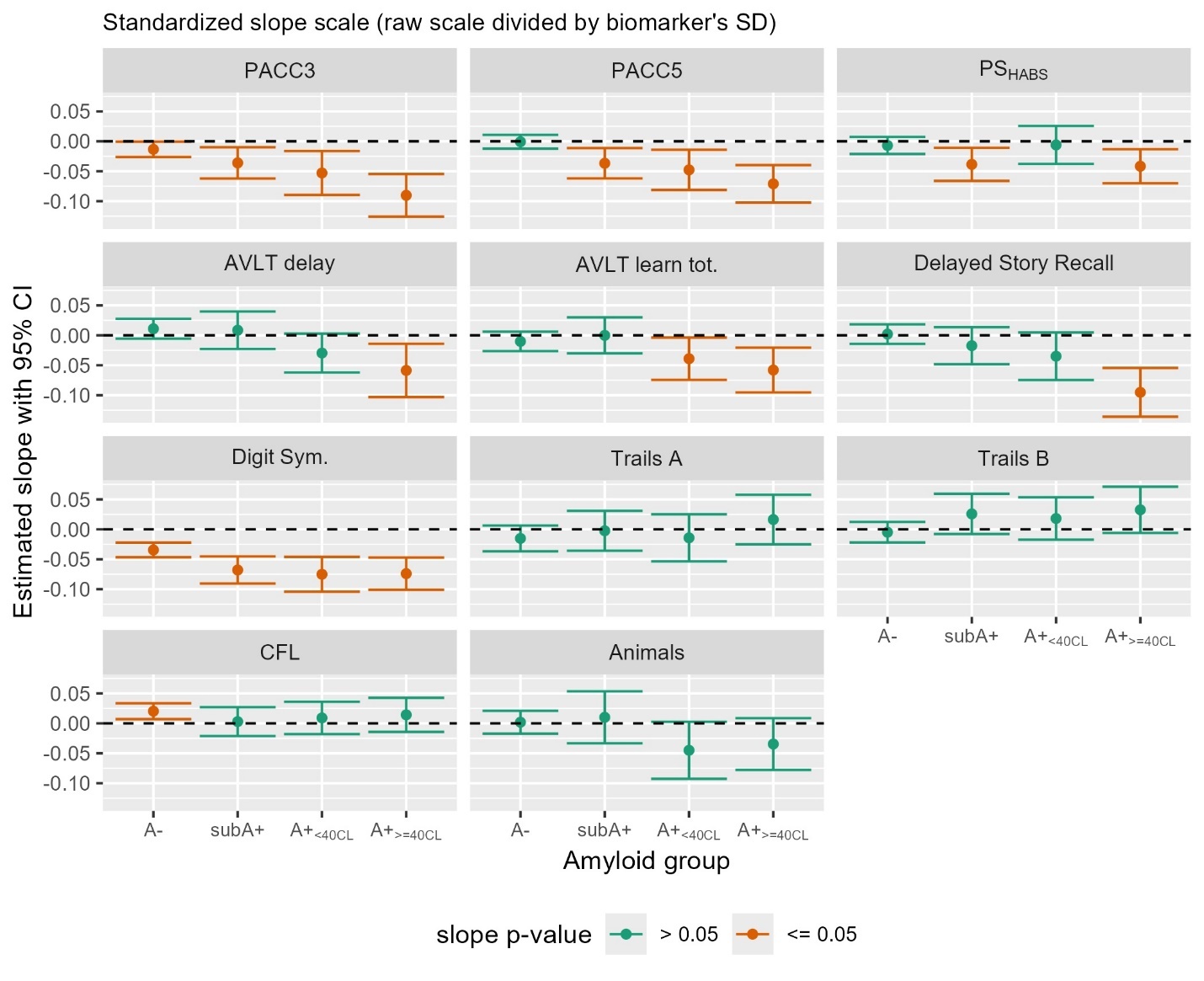

**Supplementary Figure 7 Note:** This figure is parallel to Supplementary Figures 4 and 6. In Supplementary Figure 4, the observed slopes and 95% confidence intervals (CIs) were shown using raw values from each test after subdividing the A+ group into those who are <40CL or >=40 CL. Here, those slopes and CI’s have been converted to a standardized scale (as in Supplementary Figure 6) to allow visual comparison of effect sizes across cognitive outcomes. The top row depicts the cognitive composites and the bottom three rows show individual tests, by amyloid group. Green indicates slope did not differ from 0 (p>0.05) while orange indicates slopes that differed from 0. For all but Trails A and B, negative numbers indicate decline. Abbreviations: A- = amyloid negative group (CL<~13); subA+ = sub-threshold to low positive group (CL ~13-23); and A+ = overtly A+ group (CL >~23); CL=Centiloid; Animals: Semantic fluency with animal naming; AVLT: Rey Auditory Verbal Learning Test; Digit Sym: CFL: Phonemic fluency with letters C, F, and L; CI= Confidence Interval; Digit Sym.=Digit Symbol Substitution Test; PACC: Preclinical Alzheimer’s Cognitive Composite (3 and 5 test versions); PS: Processing Speed composite (inspired by Harvard Aging Brain Study (HABS) composite).

**Supplementary Tables**
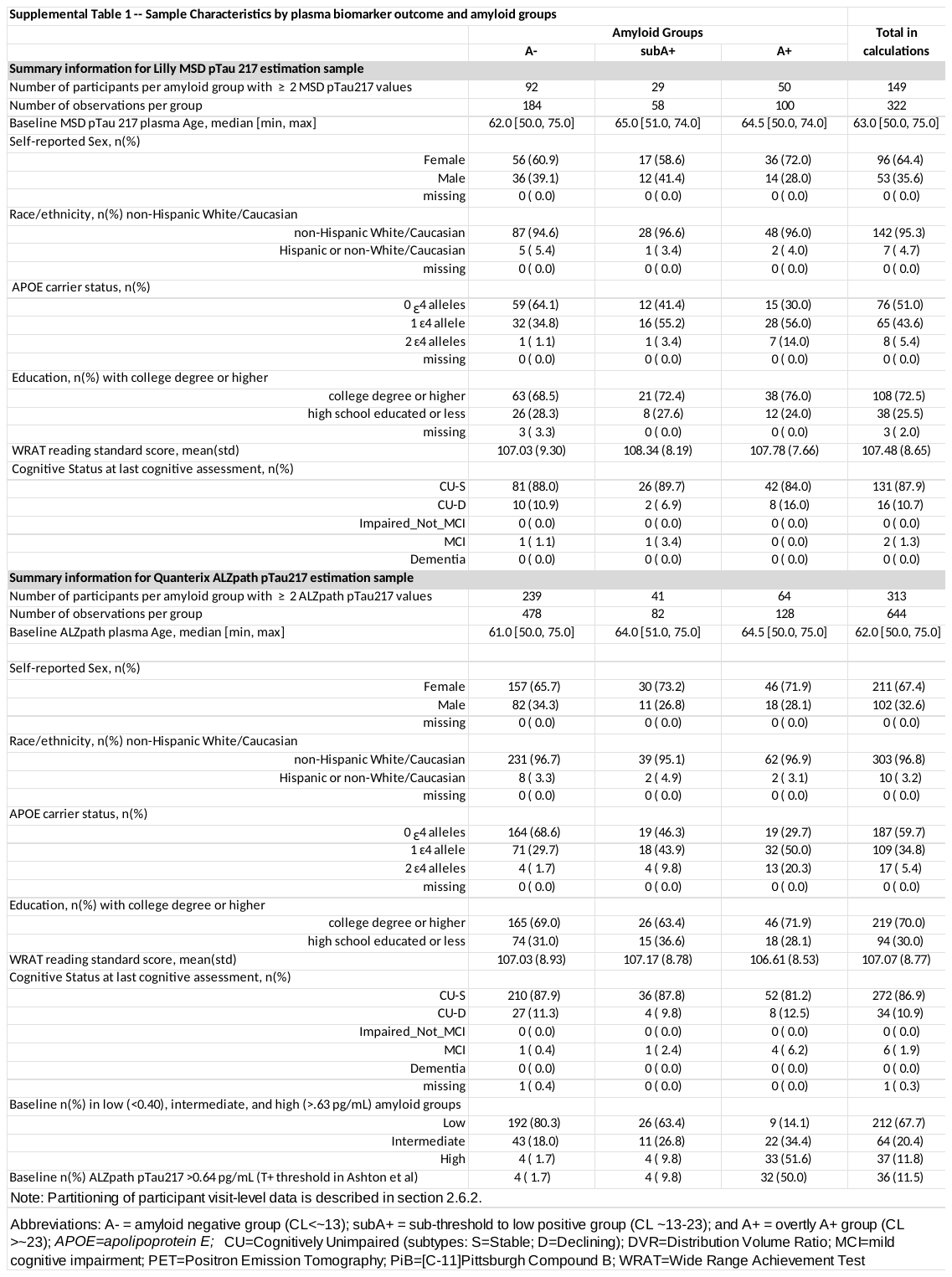

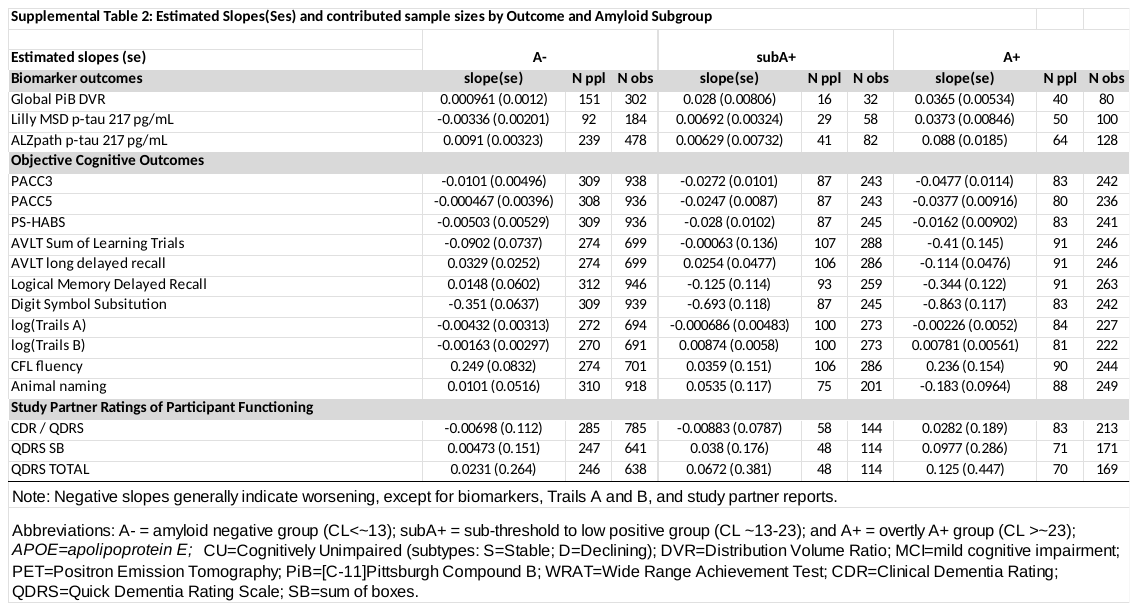

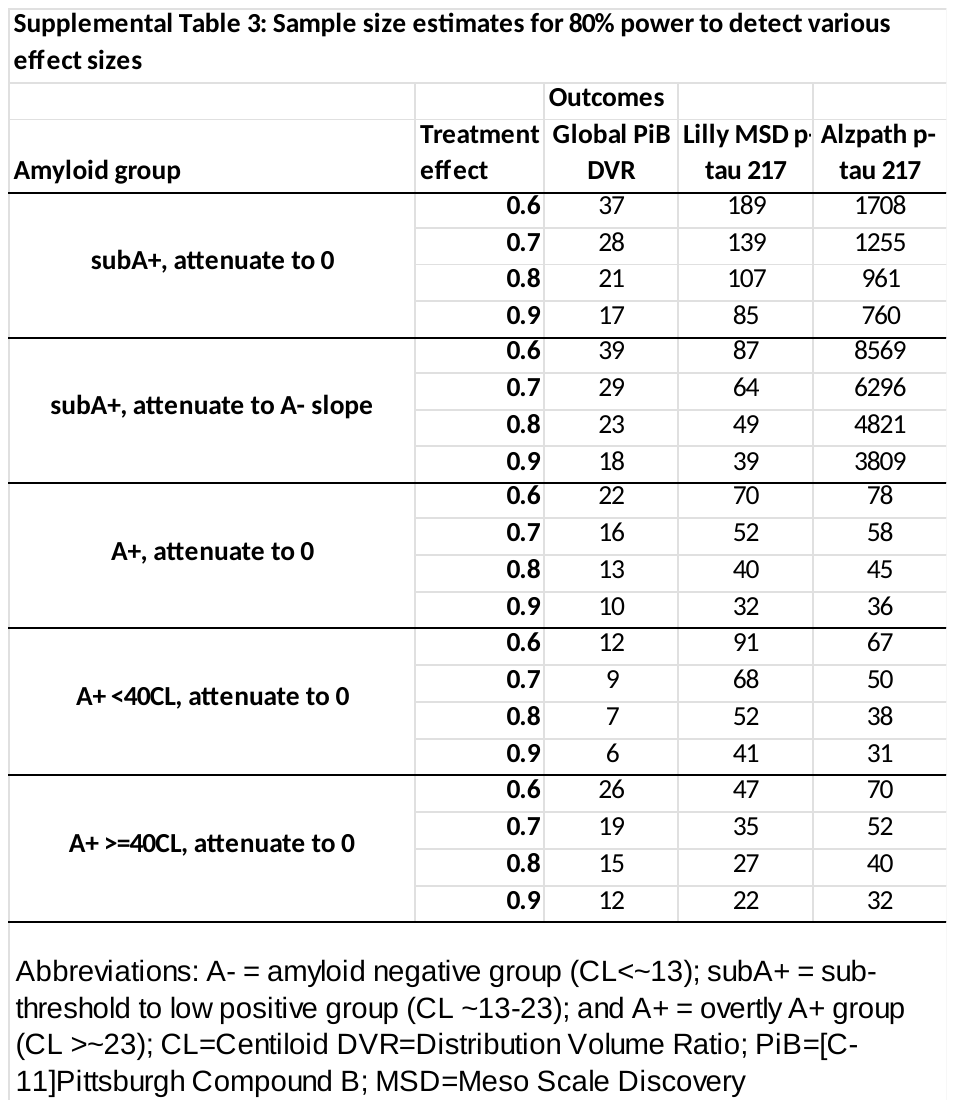

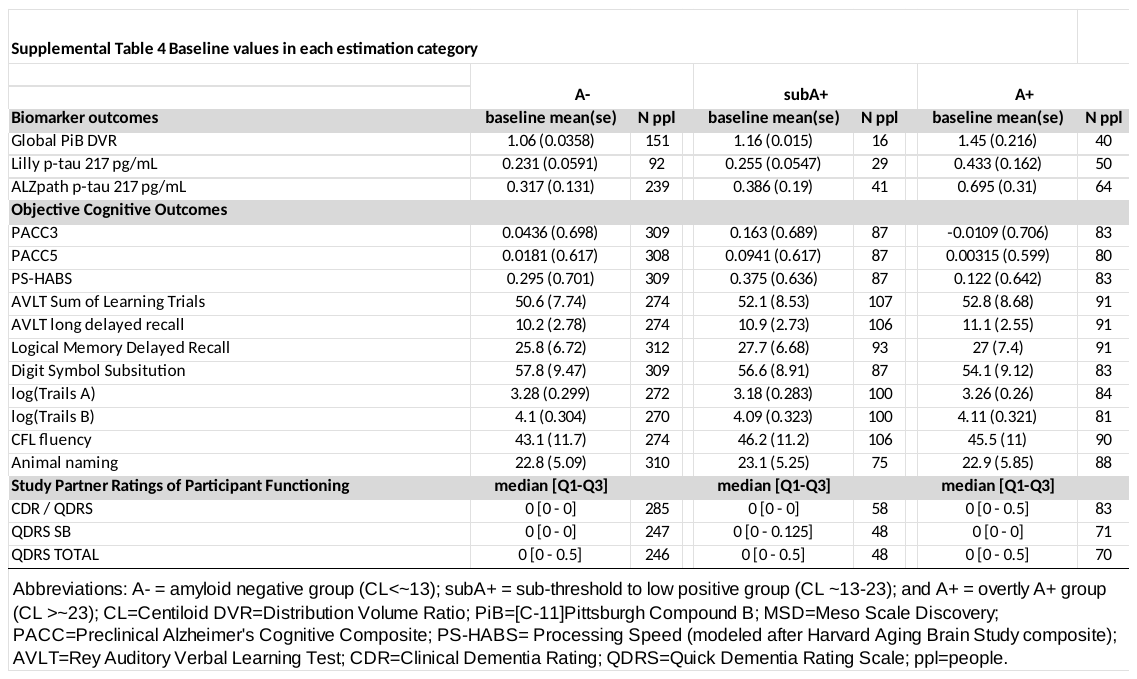

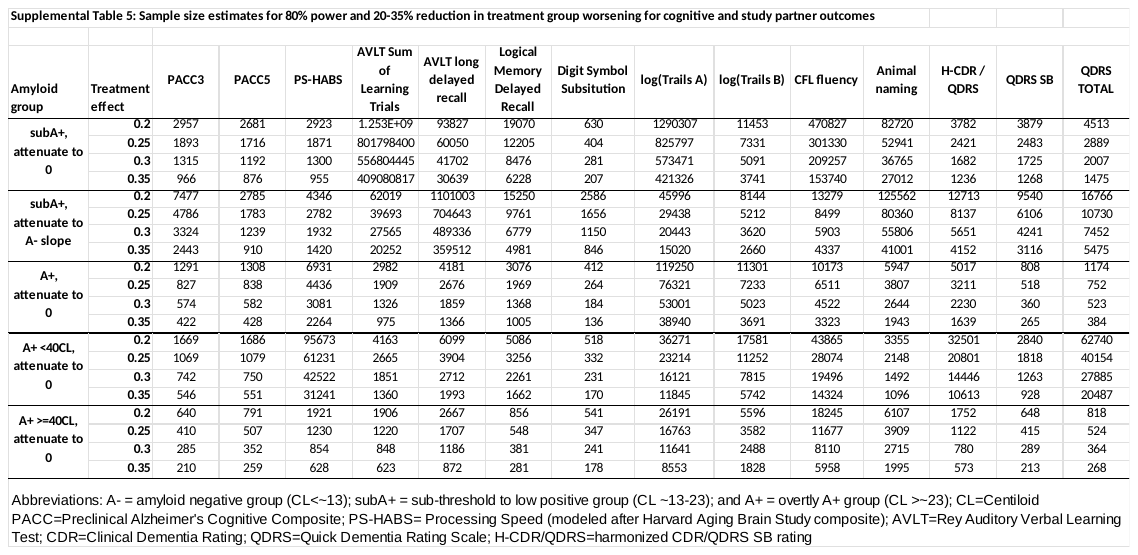
